# Do Sleep Questionnaires Add Predictive Value Beyond Demographics for Obstructive Sleep Apnea Severity? Evidence from 67,397 Patients

**DOI:** 10.64898/2026.11.01.26367019

**Authors:** Rishabh Patted, Marcus Dige, Umaer Hanif, Emmanuel Mignot, Giorgio Ricciardiello Mejia

**Author notes:** Corresponding author: Emmanuel Mignot.

## Abstract

**Background:** Sleep questionnaires are widely used to screen for obstructive sleep apnea (OSA), but their incremental value beyond collecting basic demographics is rarely assessed. Using a large sample of sleep clinic intake questionnaire data that includes the Epworth Sleepiness Scale, questions on snoring and breathing pause as well as medical history data, we quantified questionnaire-item contributions relative to demographics for OSA severity classification.

**Methods:** In a retrospective cohort of 67,397 adults referred for in-laboratory polysomnography and with questionnaire data, XGBoost classifiers were trained on all features (169 variables), questionnaire items only (161), or demographics only (age, gender, BMI, race; 4). Outcomes were the apnea–hypopnea index (AHI) and respiratory disturbance index (RDI) at 5, 15 and 30 events/h, evaluated on a held-out 10% test set; TabPFN v2.5 served as a secondary comparator. SHAP (SHapley Additive exPlanations) analysis was performed globally and within age and BMI strata; Berlin and STOP-Bang were not evaluated.

**Results:** At AHI ≥5, ≥15 and ≥30, the demographics-only model reached area under the receiver operating characteristic curve (AUROC) 0.749, 0.718 and 0.726, 0.016–0.038 below the all-features model; across all six endpoints it retained 86–94% of the above-chance AUROC. The questionnaire-only model performed worst (AUROC 0.674–0.699 for AHI). Age, BMI and gender were the leading SHAP predictors at every threshold, and the contributions of age and BMI varied across strata. Four-class severity classification was poor (XGBoost accuracy ≤0.45).

**Conclusions:** In this referred cohort, four demographic variables captured most of the achievable discrimination, and the questionnaire added at most 0.038 AUROC. Questionnaire-based screening tools should be benchmarked against a demographic baseline, and prospective validation is needed.

## 1. Introduction

Obstructive sleep apnea (OSA) is the most prevalent sleep-disordered breathing condition, affecting an estimated 936 million adults aged 30–69 years worldwide [1]. Characterized by recurrent upper-airway collapse during sleep, OSA causes intermittent hypoxia and sleep fragmentation and is associated with elevated risk of cardiovascular, metabolic, and neuropsychiatric morbidity [2,3]. Despite this burden, up to 80% of individuals with moderate-to-severe disease remain undiagnosed [4,5]. Diagnosis requires objective sleep testing: in-laboratory polysomnography (PSG) or, for uncomplicated adults at increased risk of moderate-to-severe OSA, home sleep apnea testing [6]. Both require dedicated equipment, scoring and specialist review, and access remains limited. Low-cost pre-test stratification tools are therefore needed to prioritize patients for diagnostic testing while reducing unnecessary clinical burden.

Validated sleep questionnaires—including the Epworth Sleepiness Scale (ESS) [7], the Berlin Questionnaire [8], and STOP-Bang [9]—are widely utilized as low-cost sleep apnea screening tools. In addition, sleep clinics typically use intake questionnaires that cover sleep habits, sleep apnea and other sleep disorders symptoms and basic medical history questions. Machine learning (ML) approaches applied to questionnaire-derived and demographic data have demonstrated variable discriminative performance, with most reported AUROC values in the range of 0.66–0.96 [10–21]. A critical unresolved question is whether the predictive value of these models derives primarily from demographic variables (age, gender, BMI) or from symptom information captured by the questionnaires themselves. Answering this question has direct implications for screening pathway design: if demographics dominate, extensive questionnaire administration imposes burden without proportionate gain in stratification accuracy.

In this work, we used intake questionnaire data collected by SleepMed and BioSerenity sleep laboratories between 2004 and 2019 [22] across 240 sleep disorder center facilities in 30 US states (provided in reference 22). The questionnaire contains demographic variables and over 200 questionnaire items. These ask about sleepiness, fatigue, sleep quality, sleep disturbances, comorbidities (including cardiovascular and high blood pressure) and behavioral factors. In addition, it has questions relevant to sleep apnea (snoring, breathing pauses), insomnia, parasomnia and other sleep disorders.

To address utility of questionnaire data for predicting sleep apnea, we trained and compared XGBoost classifiers under three controlled feature configurations in a cohort of 67,397 adults [22]: an All-Features model (169 variables), a Questionnaire-Only model (161 variables), and a Demographics-Only model (4 variables). Performance was evaluated across six binary classification thresholds (AHI and RDI at 5, 15, and 30 events/h), and SHAP analysis was conducted globally and within age and BMI strata to characterize the structure of feature contributions. A systematic review of prior ML-based OSA classification studies provides context for interpreting these findings. Figure 1 summarises the cohort, the study design and the main results.

**Figure 1.**
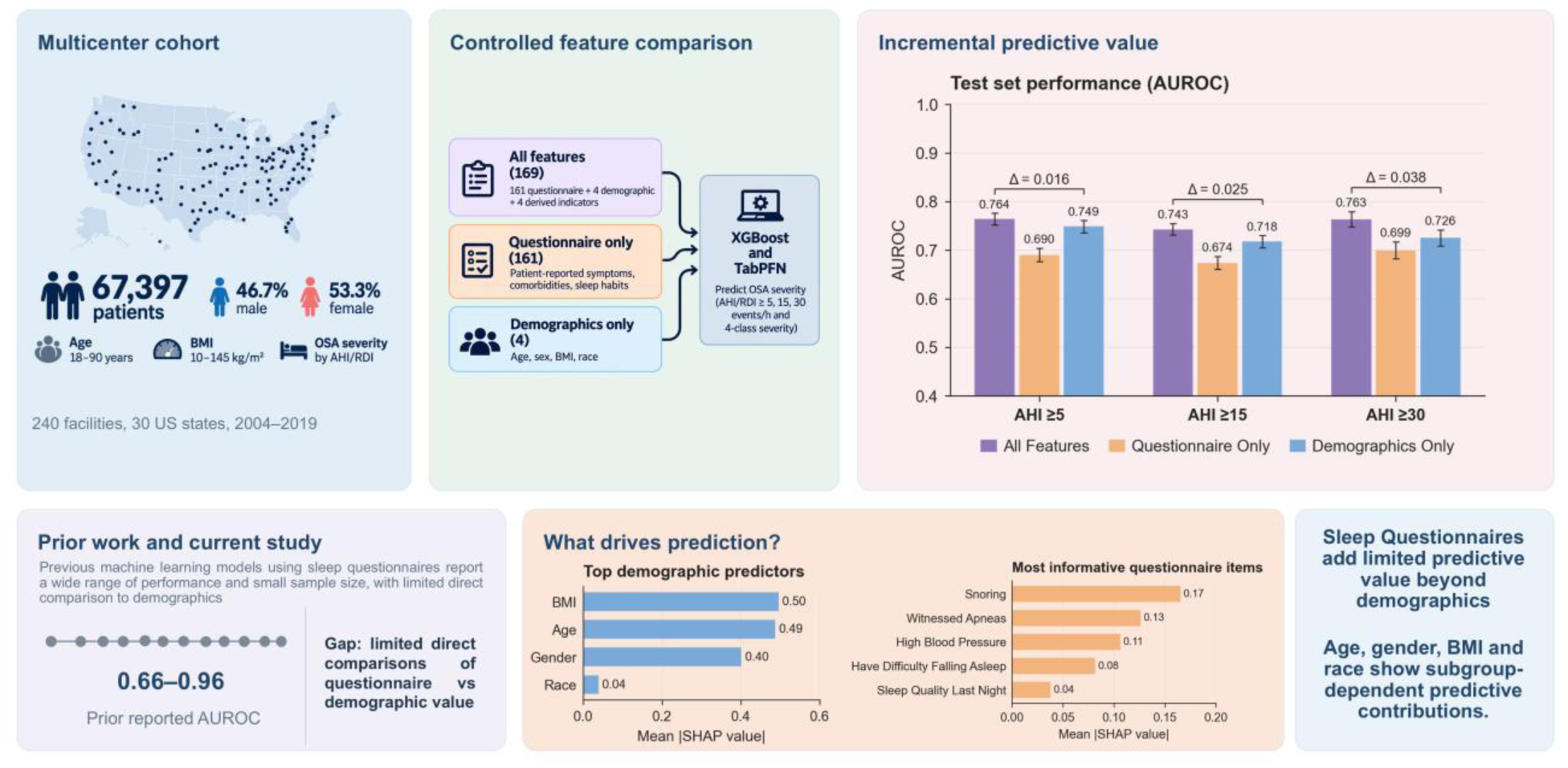
Study overview and main findings. Top: the multicenter cohort (67,397 adults, 240 facilities, 30 US states, 2004–2019; the map is schematic, dots do not mark actual facility locations), the three feature configurations compared (All-Features, 169 variables; Questionnaire-Only, 161; Demographics-Only, 4) and held-out test-set AUROC of the XGBoost models at AHI ≥5, ≥15 and ≥30 (bars, point estimate with 95% confidence interval; Δ, All-Features minus Demographics-Only AUROC by paired DeLong test). Bottom: reported AUROC of prior questionnaire-based machine-learning studies (Table 2) and mean |SHAP| in the All-Features XGBoost model, averaged over AHI ≥5, ≥15 and ≥30, for the demographic variables and the five highest-ranked questionnaire items. AHI, apnea–hypopnea index; AUROC, area under the receiver operating characteristic curve; SHAP, SHapley Additive exPlanations.

## 2. Materials and Methods

### 2.1 Study Design and Cohort

The analytic dataset is derived from a large multi-center retrospective collection comprising 540,941 records of patients referred for sleep evaluation across 240 facilities in 30 US states, with data collected by SleepMed and BioSerenity sleep laboratories between 2004 and 2019 [22]. From this resource, we identified 67,397 adults meeting inclusion criteria: a baseline diagnostic in-laboratory PSG (other study types, such as CPAP titration, split-night and multiple sleep latency studies, were excluded), age ≥18 years, no BiPAP or supplemental-oxygen use, and AHI and RDI available from recordings scored according to AASM criteria [25,27]. Patients using CPAP only were not excluded (n = 283, 0.4% of the cohort), and questionnaire items were not required to be complete. The derivation of the analytic cohort from the 540,941 source records, with the number excluded and the reason at each step, is shown in Supplementary Figure S1 and Supplementary Table S1. Studies were scored under the AASM criteria in force at each site; as the AASM revised its rules over 2004–2019, the sites adopted the updated criteria. Patient-reported data encompassed medical history, demographics, self-reported medical conditions, sleep assessment items, and medication use, collected via a sleep questionnaire designed by SleepMed that remained unchanged throughout the data collection period. PSG study outcomes were scored and physician-adjudicated at each site. Questionnaire administration occurred the night before and the night after the PSG acquisition, minimizing temporal misalignment between subjective reporting and objective assessment.

The AHI, expressed as events per hour of sleep, served as the primary measure of OSA severity and was used to define Normal (<5), Mild (5–14), Moderate (15–29), and Severe (≥30) categories.

### 2.2 Feature Configurations and Outcome Definitions

We compared three feature sets. All-Features included 169 variables: four demographic variables, four derived indicators and 161 questionnaire items. The demographic variables were age, gender, BMI and race. The four derived indicators represented underweight, normal weight, obesity, and being female and older than 50 years. The questionnaire items covered sleepiness, alertness, fatigue, sleep quality, comorbidities and behavioral factors. Questionnaire-Only included these 161 items and excluded the eight demographic and derived variables. Demographics-Only included age, gender, BMI and race. PSG-derived variables were excluded from all feature sets to prevent data leakage. The primary outcome was binary OSA classification at AHI thresholds of 5, 15, and 30 events/h; RDI-based classification at equivalent thresholds was a secondary outcome.

### 2.3 Machine Learning Pipeline

The dataset was initially partitioned into a development set (90%) and an independent held-out test set (10%) using multi-variable stratification to preserve the joint distribution of key demographic and clinical variables, including age, gender, BMI, race, self-reported snoring, witnessed apneas, laboratory site, and referring physician. The held-out test set was reserved exclusively for final evaluation. The split was drawn separately for each target, so the test sets of the six binary endpoints contain 6,763–6,802 patients; within an endpoint, all three feature sets were evaluated on the same test patients. The code is publicly available on GitHub [28].

#### 2.3.1 Gradient-Boosted Tree Models (XGBoost)

Within the 90% development set, model training and hyperparameter optimisation for XGBoost [29] were conducted without nested cross-validation: hyperparameters were selected by 5-fold multilabel-stratified cross-validation within the development set, and the selected model was evaluated once on the held-out test set. The distributions of the stratification variables and of the target classes in the 90/10 development/test split closely matched those of the full dataset (Supplementary Figures S2–S4). Hyperparameter optimisation was implemented using Optuna [30], evaluating 100 candidate configurations per target and feature set (Tree-structured Parzen Estimator sampler, seed 42). Each configuration was scored by its mean 5-fold cross-validated AUROC (binary outcomes) or macro-averaged F1 (multiclass outcomes) within the development set, and the configuration with the highest mean score was selected. The search space comprised the number of boosting rounds (100–1000), maximum tree depth (3–10), learning rate (0.01– 0.3, log scale), row subsampling (0.5–1.0), column sampling per tree (0.5–1.0), minimum child weight (1– 10), minimum split loss gamma (0–5), and L1 and L2 regularisation (1e-8 to 10, log scale). Models were trained using logistic loss for binary outcomes and softmax cross-entropy for multiclass outcomes, without early stopping. Binary models used a class weight equal to the ratio of negative to positive cases in the development set. A final model was subsequently trained on the entire 90% development set using the selected hyperparameters and evaluated once on the independent 10% held-out test set. The code applies no imputation: missing questionnaire values were passed unchanged to XGBoost and TabPFN, which handle missing values natively, and the missingness-indicator columns were not model features. Missingness was 29.5% for snoring, 32.4% for witnessed apneas and 20.1% for high blood pressure; age, gender, BMI and race were complete.

#### 2.3.2 TabPFN Models

TabPFN v2.5 [31, 32], a transformer-based foundation model for tabular data, was evaluated as a secondary comparator under a distinct framework consistent with its design constraints. As TabPFN does not require task-specific hyperparameter tuning, no nested cross-validation or optimisation procedure was applied. Instead, TabPFN was evaluated using 5-fold stratified cross-validation across the full dataset without a separate held-out test set. In each fold, 80% of data served as the conditioning set and the remaining 20% as the evaluation set. The default TabPFN v2.5 checkpoint was used (ModelVersion.V2_5).

#### 2.3.3 Performance Evaluation and Thresholding

All performance metrics (AUROC, sensitivity, specificity, precision, and F1 score) were computed on held-out data: the 10% test set for XGBoost and out-of-fold predictions for TabPFN. Operating thresholds for binary classification were fixed at τ=0.50 for XGBoost and TabPFN. F1 was computed for the positive class and equals the harmonic mean of precision and sensitivity. Four-class predictions were assigned to the class with the highest predicted probability (argmax), so the 0.50 threshold applies only to the binary models. For XGBoost, sensitivity, specificity, precision and F1 were computed from the held-out test-set predictions at this threshold; for TabPFN they were computed within each cross-validation fold and are reported as mean ± SD across the five folds. Approximate 95% confidence intervals for AUROC (Hanley– McNeil), for AUROC differences, and Wilson intervals for sensitivity, specificity and precision were computed post hoc from the reported point estimates and the exact numbers of cases and controls in each endpoint’s test set (Supplementary Tables S2 and S3); paired DeLong tests were not possible because per-patient predictions were not available for this analysis. To assess clinical utility beyond discrimination, decision-curve analysis compared net benefit between feature sets across threshold probabilities 0.05–0.50 on the XGBoost held-out test set, with patient-level stratified bootstrap 95% confidence intervals for paired differences at thresholds 0.10, 0.20 and 0.30 (1,000 resamples). Calibration of the binary XGBoost probabilities was assessed via logistic recalibration (slope and intercept) and Brier score on the held-out test set; probabilities were not recalibrated before use in the decision-curve analysis.

#### 2.3.4 Model Interpretability

Model interpretability was assessed using SHAP for XGBoost models, with feature attribution analyses conducted on the final trained model. Stratified SHAP analyses were performed across predefined age groups (≤36, 36–46, 46–58, ≥58 years) and BMI categories (≤26, 26–30, 30–34, ≥34 kg/m²) to evaluate heterogeneity in feature importance across clinically relevant subpopulations. SHAP-based interpretability was not applied to TabPFN given its probabilistic inference structure.

## 3. Results

### 3.1 Cohort Characteristics

A total of 67,397 unique subjects with mean age 54.3 ± 15.6 years and 46.7% males were utilized in the analytical dataset (Table 1; full characteristics in Supplementary Table S4). Across OSA severity clusters, age increases progressively, with mean age rising from 48.6 ± 16.3 years in the normal group to 59.1 ± 14.1 years in the severe group, indicating a clear age gradient. The proportion of females declines markedly with severity, from 62.3% in normal to 36.6% in severe, suggesting a shift toward male predominance in higher OSA burden. Median age follows the same trend (48.0 [36.0–61.0] vs. 61.0 [49.0–70.0]).

**Table 1.**
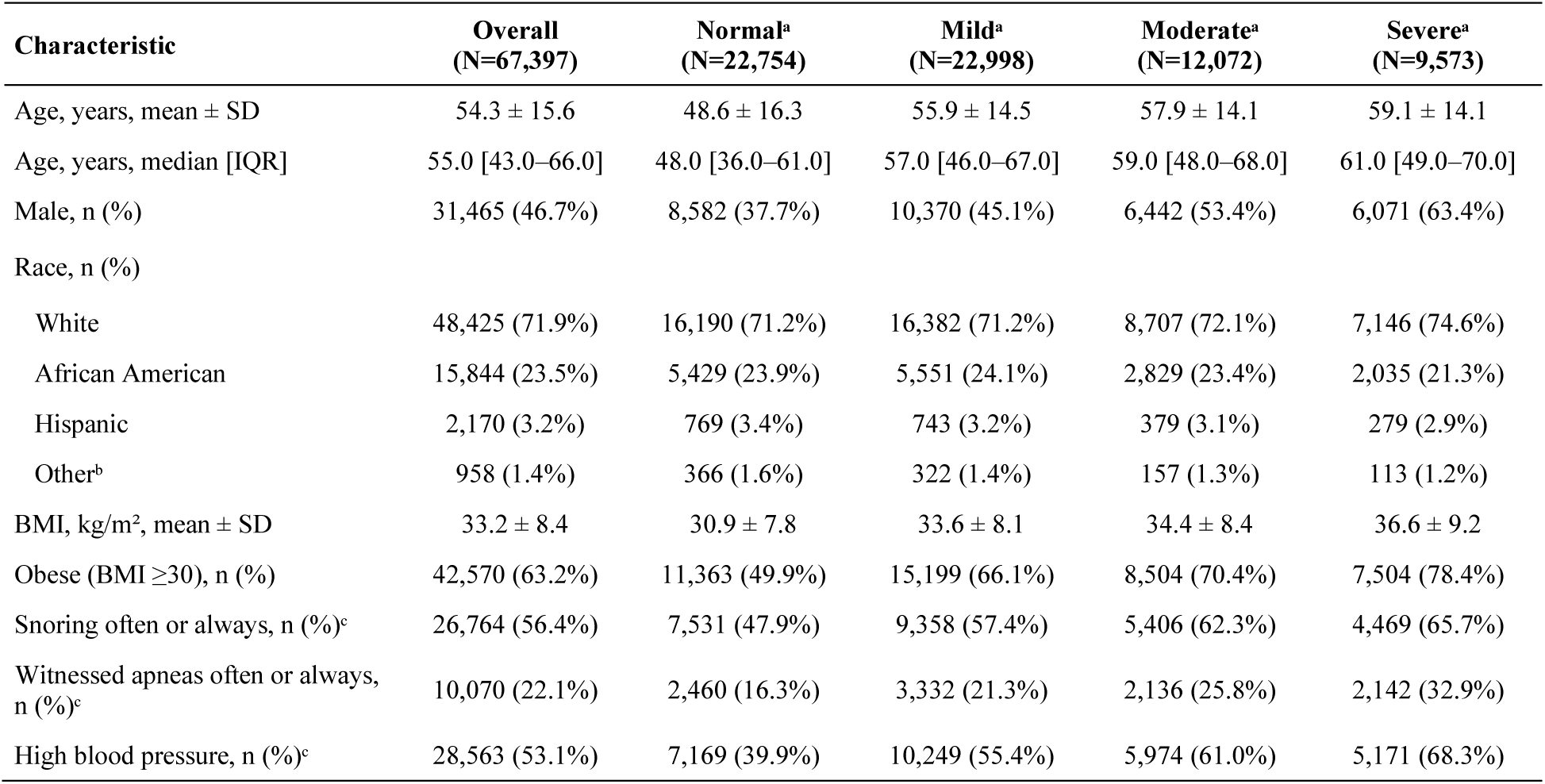
Characteristics of the analytical cohort by OSA severity (AHI, events/h). Values are mean ± SD, median [IQR] or n (%). AHI, apnea–hypopnea index; BMI, body mass index; IQR, interquartile range. ^a^Normal, AHI <5; Mild, 5–14; Moderate, 15–29; Severe, ≥30 events/h. _b_American Indian, Pacific Islander, Asian and Alaska Native. ^c^Percentage of participants with a non-missing response. Full characteristics are given in Supplementary Table S4.

### 3.2 Comparison with existing literature

To compare our results with current evidence, we conducted a systematic review of ML approaches for OSA classification using questionnaire-derived features, following PRISMA guidance [23,24]. The search spanned 2016–2025 and included Google Scholar, PubMed, and Embase. After screening 837 records, 12 studies met inclusion criteria (PSG-based AHI as reference standard; reported sensitivity, specificity, and/or AUROC; adult populations). Key characteristics are summarized in Table 2; PRISMA flow, QUADAS-2 risk-of-bias assessment, and summary ROC analyses are provided in Supplementary Figures S5–S7 and Table S5.

**Table 2.**
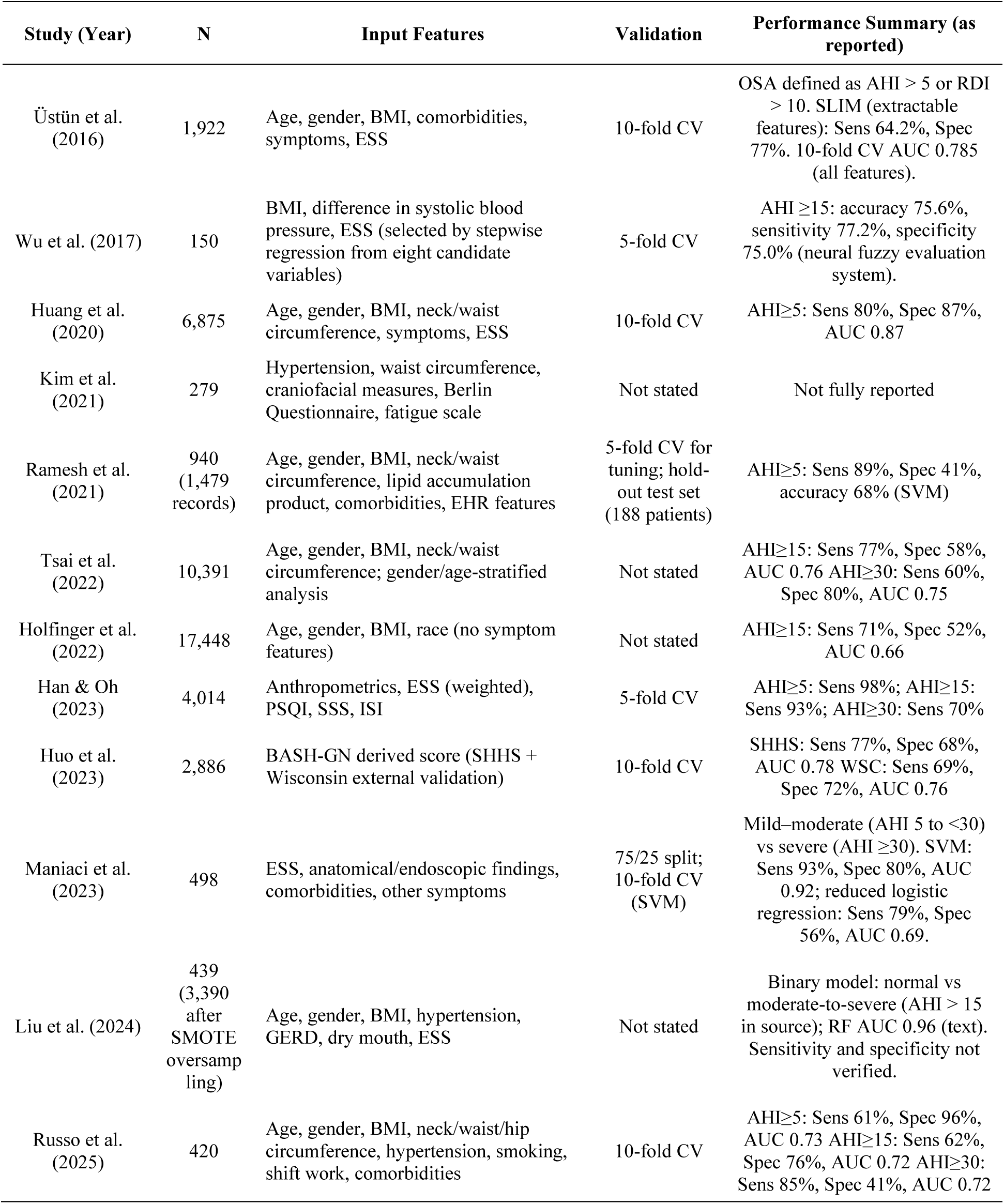
State-of-the-art studies using questionnaire-derived features for ML-based OSA classification. AHI, apnea– hypopnea index; AUC, area under ROC curve; BMI, body mass index; ESS, Epworth Sleepiness Scale; Sens, sensitivity; Spec, specificity. Performance values are as reported in source tables.

| **Study (Year)** | **N** | **Input Features** | **Validation** | **Performance Summary (as reported)** |
|---|---|---|---|---|
| Üstün et al. (2016) | 1,922 | Age, gender, BMI, comorbidities, symptoms, ESS | 10-fold CV | OSA defined as AHI > 5 or RDI > 10. SLIM (extractable features): Sens 64.2%, Spec 77%. 10-fold CV AUC 0.785 (all features). |
| Wu et al. (2017) | 150 | BMI, difference in systolic blood pressure, ESS (selected by stepwise regression from eight candidate variables) | 5-fold CV | AHI ≥15: accuracy 75.6%, sensitivity 77.2%, specificity 75.0% (neural fuzzy evaluation system). |
| Huang et al. (2020) | 6,875 | Age, gender, BMI, neck/waist circumference, symptoms, ESS | 10-fold CV | AHI≥5: Sens 80%, Spec 87%, AUC 0.87 |
| Kim et al. (2021) | 279 | Hypertension, waist circumference, craniofacial measures, Berlin Questionnaire, fatigue scale | Not stated | Not fully reported |
| Ramesh et al. (2021) | 940 (1,479 records) | Age, gender, BMI, neck/waist circumference, lipid accumulation product, comorbidities, EHR features | 5-fold CV for tuning; hold-out test set (188 patients) | AHI≥5: Sens 89%, Spec 41%, accuracy 68% (SVM) |
| Tsai et al. (2022) | 10,391 | Age, gender, BMI, neck/waist circumference; gender/age-stratified analysis | Not stated | AHI≥15: Sens 77%, Spec 58%, AUC 0.76 AHI≥30: Sens 60%, Spec 80%, AUC 0.75 |
| Holfinger et al. (2022) | 17,448 | Age, gender, BMI, race (no symptom features) | Not stated | AHI≥15: Sens 71%, Spec 52%, AUC 0.66 |
| Han & Oh (2023) | 4,014 | Anthropometrics, ESS (weighted), PSQI, SSS, ISI | 5-fold CV | AHI≥5: Sens 98%; AHI≥15: Sens 93%; AHI≥30: Sens 70% |
| Huo et al. (2023) | 2,886 | BASH-GN derived score (SHHS + Wisconsin external validation) | 10-fold CV | SHHS: Sens 77%, Spec 68%, AUC 0.78 WSC: Sens 69%, Spec 72%, AUC 0.76 |
| Maniaci et al. (2023) | 498 | ESS, anatomical/endoscopic findings, comorbidities, other symptoms | 75/25 split; 10-fold CV (SVM) | Mild–moderate (AHI 5 to <30) vs severe (AHI ≥30). SVM: Sens 93%, Spec 80%, AUC 0.92; reduced logistic regression: Sens 79%, Spec 56%, AUC 0.69. |
| Liu et al. (2024) | 439 (3,390 after SMOTE oversampling) | Age, gender, BMI, hypertension, GERD, dry mouth, ESS | Not stated | Binary model: normal vs moderate-to-severe (AHI > 15 in source); RF AUC 0.96 (text). Sensitivity and specificity not verified. |
| Russo et al. (2025) | 420 | Age, gender, BMI, neck/waist/hip circumference, hypertension, smoking, shift work, comorbidities | 10-fold CV | AHI≥5: Sens 61%, Spec 96%, AUC 0.73 AHI≥15: Sens 62%, Spec 76%, AUC 0.72 AHI≥30: Sens 85%, Spec 41%, AUC 0.72 |

Across all included studies, the most consistently reported predictors were age, gender, BMI, and neck/waist circumference, frequently augmented by snoring, witnessed apneas, and ESS scores. Predictive performance was heterogeneous across AHI thresholds, with reported AUROC values varying according to disease severity definitions, population characteristics, and validation strategies. Summary ROC meta-analysis (Moses–Littenberg sROC; 4, 6 and 3 study cohorts) yielded diagnostic odds ratios (DOR) of 13.6, 5.4 and 4.8 at AHI thresholds of 5, 15 and 30 events/h, respectively (Supplementary Table S6); with so few cohorts per threshold these estimates are imprecise. Concentrated risk of bias in patient selection and index-test domains, alongside inconsistent reporting of validation strategies, limits the interpretability and generalizability of prior evidence.

### 3.3 Model Performance

#### 3.3.1 XGBoost

Across all severity thresholds, models incorporating the full feature set achieved the highest discrimination (Figure 2). For AHI-based classification, AUROC ranged from 0.743–0.764 with All-Features compared to 0.718–0.749 for Demographics-Only, while Questionnaire-Only models consistently underperformed (AUROC 0.674–0.699). A similar pattern was observed for RDI thresholds, where All-Features models reached AUROCs of 0.746–0.763 versus 0.717–0.746 for Demographics-Only and 0.679–0.696 for Questionnaire-Only (Table 3; Supplementary Table S7). The largest AUROC difference between All-Features and Demographics-Only was 0.038. Removing the four demographic variables and four derived indicators reduced AUROC by 0.062–0.074 across all six endpoints. These estimates came from a single held-out test split. The approximate, conservative 95% confidence intervals excluded zero for the All-Features minus Questionnaire-Only difference at every endpoint, and for the All-Features minus Demographics-Only difference at four of the six endpoints; at AHI ≥5 and RDI ≥5 the lower bound was approximately zero. Supplementary Table S2 reports the confidence intervals.

**Figure 2.**
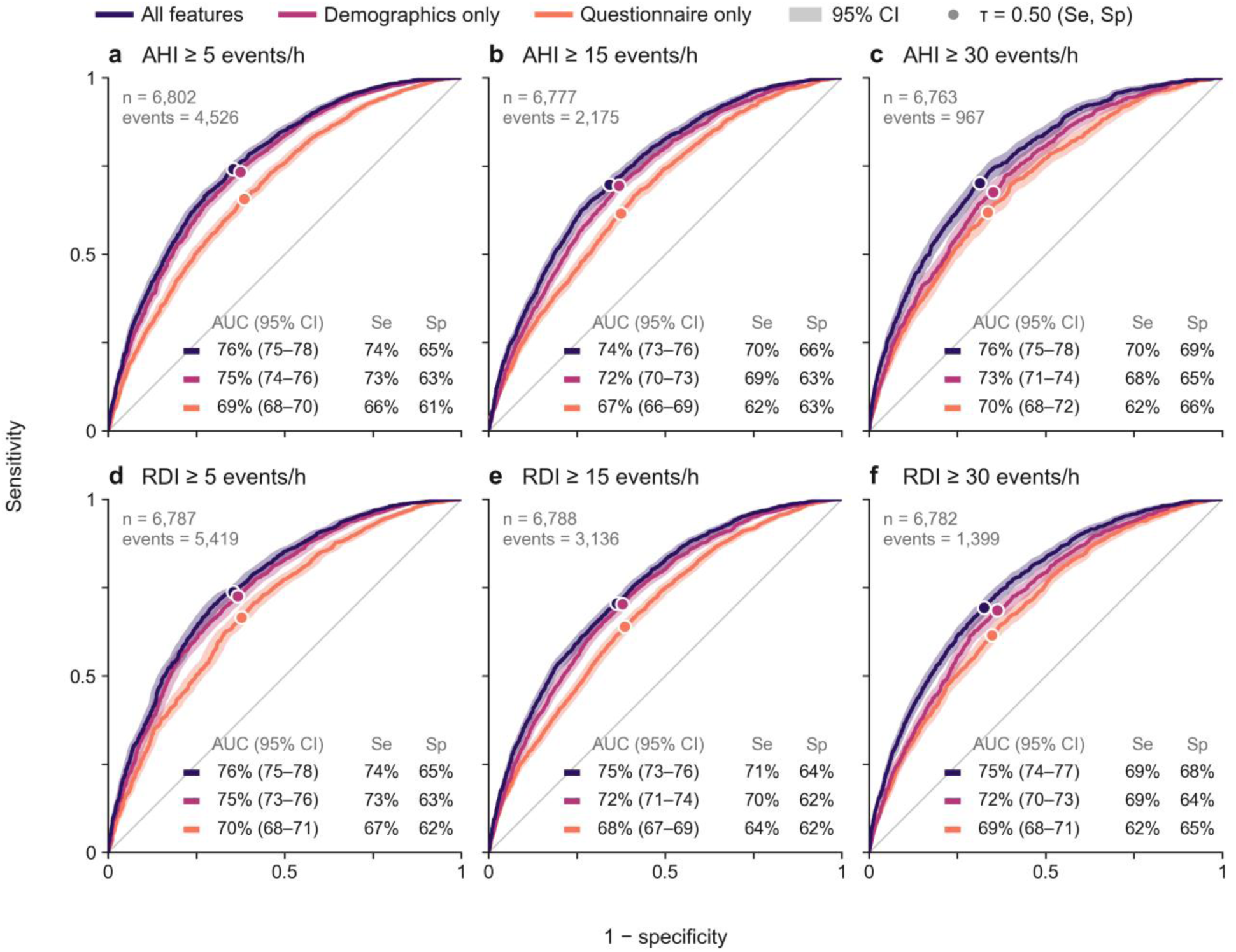
Receiver operating characteristic (ROC) curves for the XGBoost models on the held-out test set for the apnea–hypopnea index (AHI ≥5, ≥15, ≥30; panels a–c) and respiratory disturbance index (RDI ≥5, ≥15, ≥30; panels d–f). Each row is one feature set, with its AUROC (AUC) and its sensitivity (Se) and specificity (Sp) at the fixed 0.50 threshold. All-Features models (demographics + questionnaire) achieved the highest discrimination at every threshold, followed by Demographics-Only and then Questionnaire-Only models.

**Table 3.**
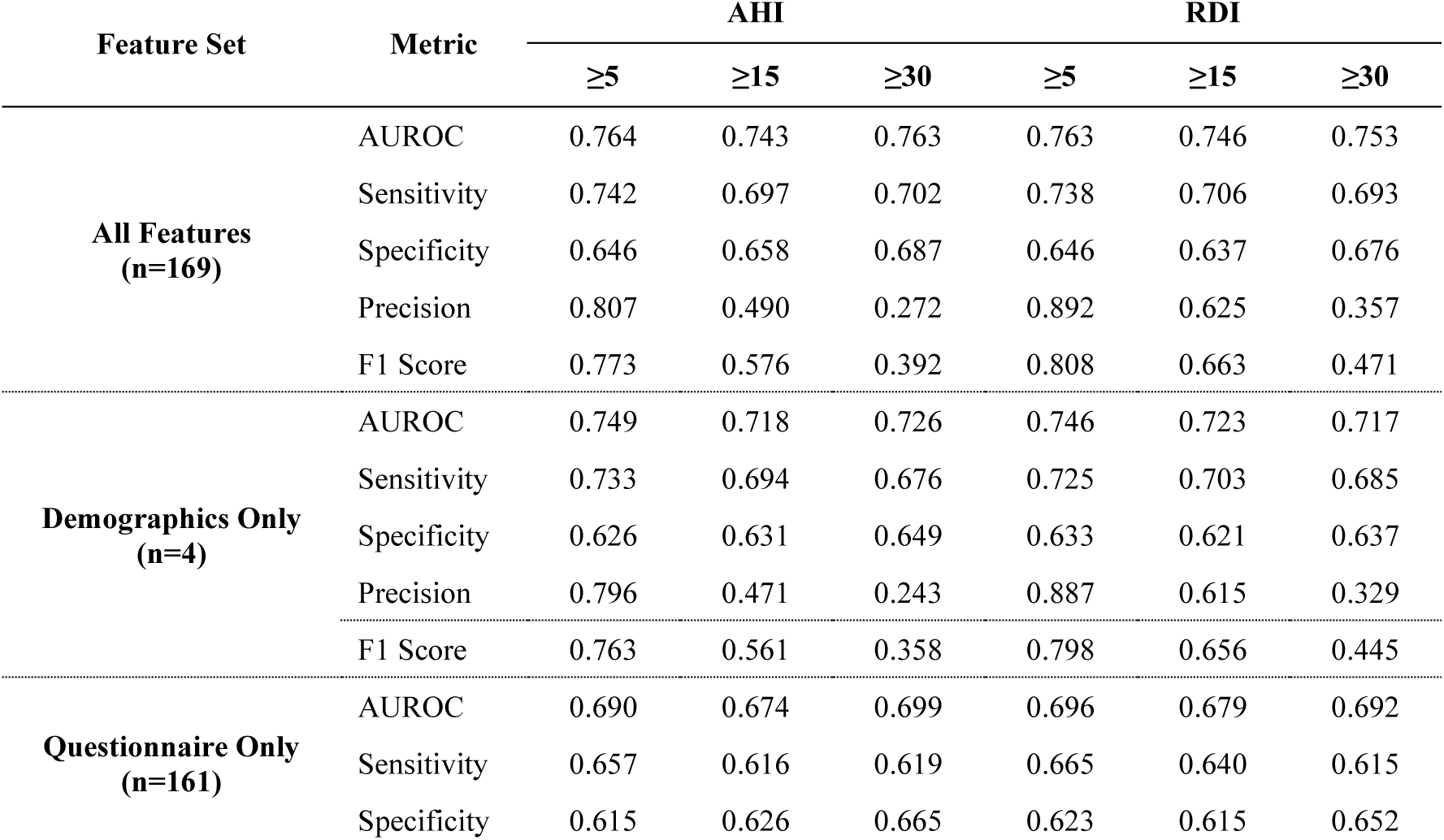
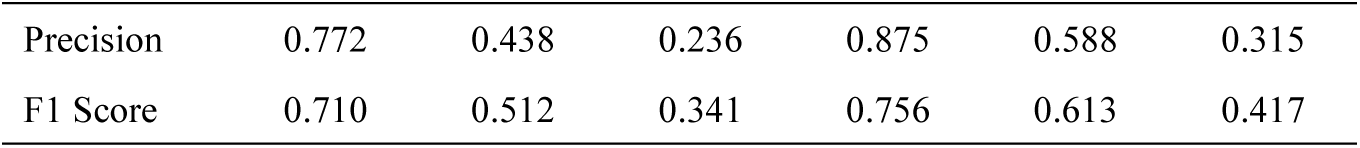
Predictive performance of XGBoost models for binary classification across feature sets and diagnostic thresholds (held-out test set; fixed decision threshold 0.50). Within each endpoint the three feature sets were evaluated on the same test patients (6,763–6,802 per endpoint). F1 is computed for the positive class from the same predictions and equals the harmonic mean of precision and sensitivity. AUROC, area under the receiver operating characteristic curve; AHI, apnea–hypopnea index; RDI, respiratory disturbance index.

Decision-curve analysis showed that the All-Features model yielded a small positive net-benefit gain over Demographics-Only and Questionnaire-Only models at clinically plausible thresholds (0.10–0.30), most clearly at the higher AHI and RDI cut-offs: at AHI ≥30, the All-Features minus Demographics-Only difference in net benefit was 0.0045–0.0318 and the All-Features minus Questionnaire-Only difference was 0.0071–0.0591 across thresholds 0.10–0.30, with 95% bootstrap intervals excluding zero; gains were smaller and less consistent at AHI ≥5 and RDI ≥5 (Supplementary Table S8, Figure S8). Calibration slopes were close to 1 (0.93–1.05) but intercepts were offset, consistent with the class weighting used in training; probabilities were not recalibrated, so these net-benefit estimates are provisional (Supplementary Table S9).

Four-class classification (None, Mild, Moderate, Severe) was substandard for every feature set. Predictions were assigned to the class with the highest predicted probability. Macro-averaged F1 was 0.32–0.40 and accuracy 0.37–0.45, against majority-class accuracies of 0.341 (AHI) and 0.339 (RDI) (Supplementary Figure S9, Tables S2 and S10). Moderate disease was rarely identified: for All-Features, recall for the Moderate class was 0.09 (AHI) and 0.20 (RDI), and most moderate cases were predicted as Mild. SHAP attributions for the four-class models showed age, BMI, and gender among the most influential predictors for most severity classes, consistently so for RDI; for the AHI Mild class, the leading predictors were difficulty falling asleep, age and BMI, with gender outside the top ten (Supplementary Figures S10 and S11, Table S11). On the four-class task, All-Features performed modestly better than Demographics-Only on all four reported metrics. Both performed better than Questionnaire-Only. Table S10 gives the macro-F1 and accuracy values for AHI and RDI.

#### 3.3.2 TabPFN

TabPFN demonstrated comparable binary AUROC performance to XGBoost across endpoints (AUROC range: 0.742–0.773), but exhibited fundamentally different operating behaviours across severity thresholds (Supplementary Table S12).

For AHI-based classification (Figure 3), performance was characterised by a pronounced shift from high-recall/low-specificity at low thresholds to high-specificity/low-recall at high thresholds. At AHI ≥5, the model achieved strong recall (0.901 ± 0.004) but poor specificity (0.433 ± 0.005), indicating systematic over-classification. This pattern inverted at AHI ≥30, where specificity became near-perfect (0.989 ± 0.001) but recall collapsed (0.096 ± 0.010), yielding a markedly low F1 of 0.166 ± 0.016. AHI ≥15 represented a transition regime, with high specificity (0.896 ± 0.006) but substantially reduced recall (0.341 ± 0.006). Relative to majority-class accuracy, TabPFN accuracy was higher by 0.081, 0.039 and 0.004 at AHI ≥5, ≥15 and ≥30, and by 0.024, 0.133 and 0.011 at RDI ≥5, ≥15 and ≥30 (Supplementary Table S2); at AHI ≥30 the accuracy of 0.862 is therefore essentially the majority-class rate (0.858).

**Figure 3.**
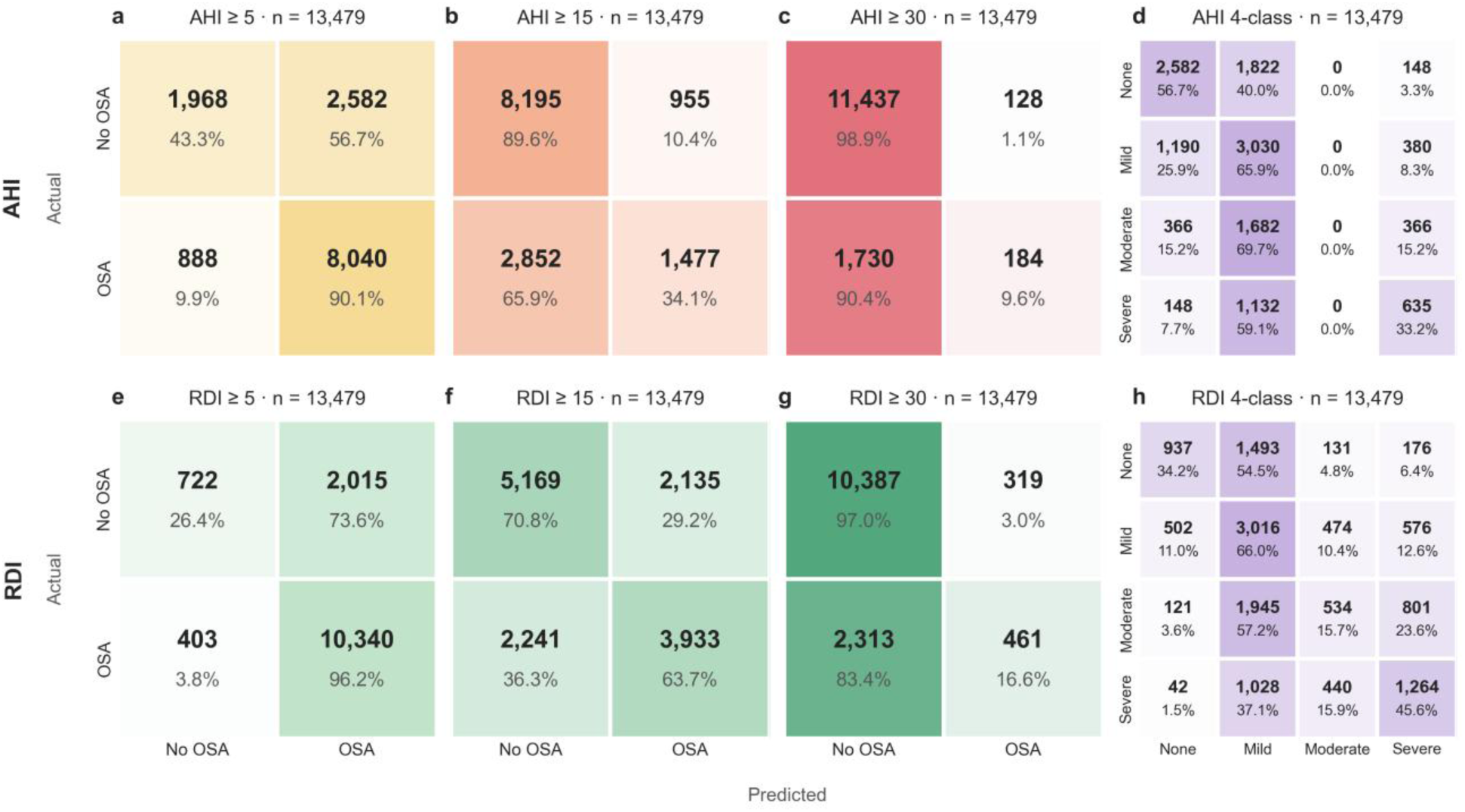
Confusion matrices showing mean TabPFN performance across the five cross-validation folds for OSA classification (cell counts averaged across folds; n = 13,479 per fold). Binary predictions used a fixed decision threshold of 0.50. Panels (a–c) show binary classification at AHI ≥5, ≥15 and ≥30, and panel (d) four-class severity classification for AHI. Panels (e–g) show the corresponding RDI thresholds, and panel (h) four-class severity classification for RDI; four-class predictions were assigned to the class with the highest predicted probability. Each cell gives the count and, beneath it, the percentage of the actual class (row); shading deepens with that percentage.

RDI-based models demonstrated more balanced sensitivity–specificity trade-offs at intermediate thresholds, particularly at RDI ≥15 (recall 0.637 ± 0.007; specificity 0.708 ± 0.004; F1 0.643 ± 0.006). These patterns are consistent with the fixed 0.50 cutoff yielding asymmetric operating points as class prevalence changes across thresholds, rather than with a limitation in rank discrimination. Four-class severity classification yielded modest macro-averaged F1 scores (AHI: 0.362 ± 0.004; RDI: 0.400 ± 0.004). For AHI, no patient was predicted as Moderate and 69.7% of moderate cases were predicted as Mild (Figure 3d); for RDI, 57.2% of moderate cases were predicted as Mild (Figure 3h). This pattern suggests that demographic and questionnaire features separate the adjacent intermediate severity classes poorly.

### 3.4 SHAP Feature Importance

SHAP feature importance was evaluated in the trained XGBoost All-Features model (Figure 4). Age, BMI, and gender consistently emerged as the dominant predictors across all AHI thresholds, with a descriptive shift in relative importance as severity increased.

**Figure 4.**
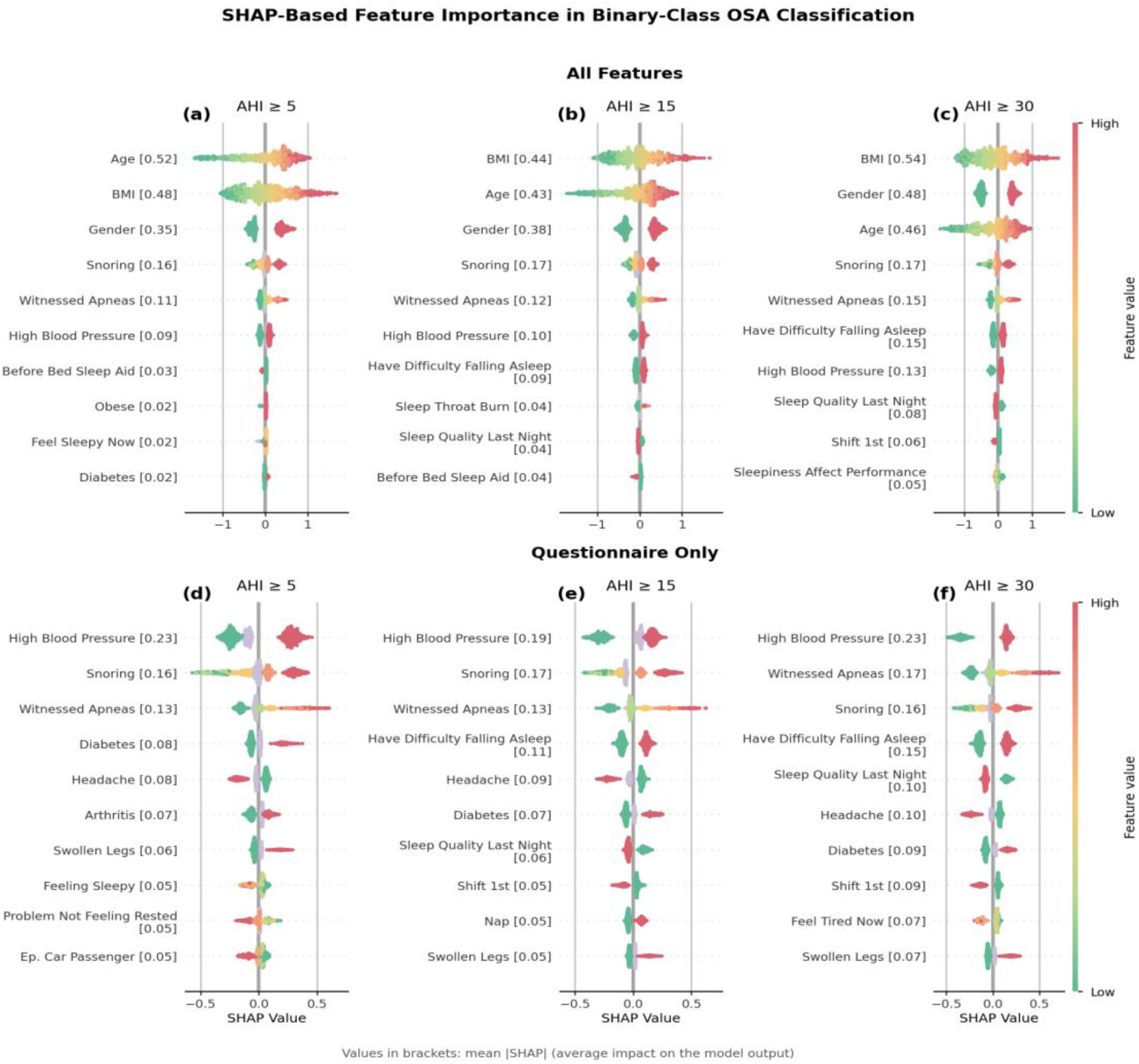
SHAP-based feature importance for binary classification of OSA across three AHI thresholds (≥5, ≥15, ≥30 events/h). Panels (a–c) display results for the All-Features model; panels (d–f) show the Questionnaire-Only model. Features are ranked by mean absolute SHAP value (value in brackets, log-odds), and the SHAP axis is shared within each row; points represent individual observations colored by feature value (green = low, red = high; lavender = missing).

In the All-Features model, age ranked highest at AHI ≥5 (mean |SHAP| 0.527) and BMI at AHI ≥30 (0.543); age and BMI were nearly tied at AHI ≥15 (0.455 vs 0.448). Gender’s mean |SHAP| rose from 0.351 to 0.476 across thresholds. The ordering of the top two features changes across thresholds, but the differences are small and based on single-model point estimates without uncertainty. Together, age, BMI and gender accounted for 51.1%, 40.5% and 39.1% of the summed mean |SHAP| over all 169 features at AHI ≥5, ≥15 and ≥30, respectively, and the feature rankings were positively correlated across endpoints (Spearman ρ 0.61–0.86; Supplementary Table S13), so all remaining features contributed substantially less individually (Supplementary Table S14).

In the Questionnaire-Only model (Figure 4d–f), high blood pressure was the top-ranked predictor across all thresholds (mean |SHAP| 0.19–0.23). Snoring and witnessed apneas were the only questionnaire items with consistent face validity as direct indicators of nocturnal respiratory disturbance. Subjective sleepiness-related variables demonstrated minimal or inconsistent contributions across all thresholds, indicating limited utility for stratifying OSA severity.

### 3.5 Stratified SHAP Analysis

Stratified analyses at AHI ≥5 showed that the ranking of features changed across age and BMI strata (Figure 5). Features were ranked separately within each stratum, so each panel carries its own feature labels. In the youngest age group (≤36 years), age ranked first, but its SHAP values were almost all negative (about −1.7 to −0.4), so its rank mainly reflects a near-constant downward shift; BMI ranked second and had the widest spread of SHAP values (about −1.1 to +1.6; Figure 5a). In the two middle age groups (>36 to ≤46 and >46 to ≤58 years), BMI ranked first (Figure 5b, 5c). In the oldest group (>58 years), age ranked first again and BMI retained a wide spread of SHAP values (Figure 5d). Gender contributions did not increase with age; they were smaller in the >58 group (about −0.3 to +0.5) than in the ≤36 group (about −0.45 to +0.7). Within each age stratum the stratifying variable has a restricted range, so its own SHAP spread and rank are not directly comparable with those of the other strata.

**Figure 5.**
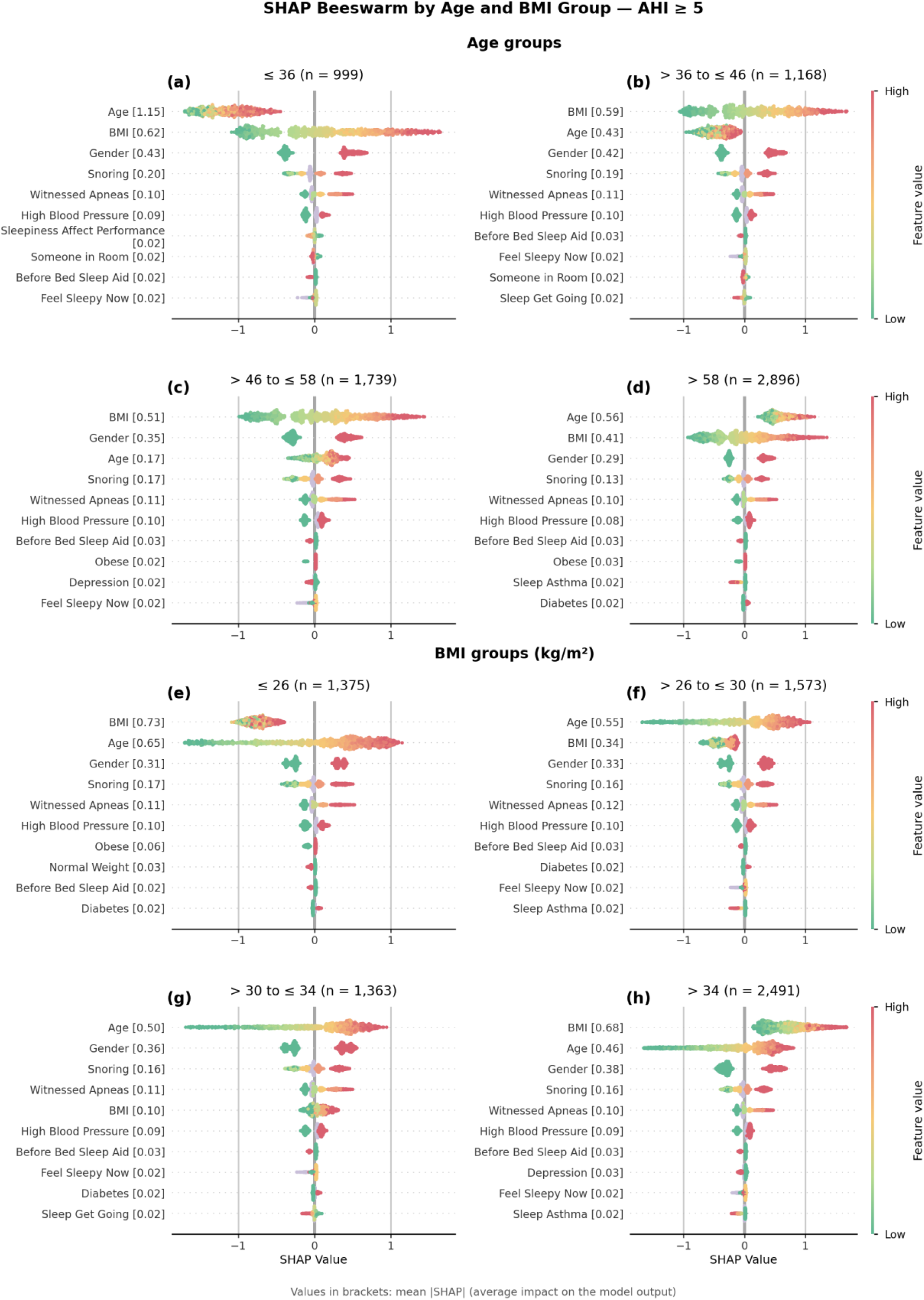
SHAP summary plots for the All-Features XGBoost binary model at AHI ≥5 (test set), stratified by age group (top; ≤ 36, > 36 to ≤ 46, > 46 to ≤ 58, > 58 years; panels a–d) and by BMI category (bottom; ≤ 26, > 26 to ≤ 30, > 30 to ≤ 34, > 34 kg/m²; panels e–h). With n the number of test-set patients in the stratum. Features were ranked by mean absolute SHAP value separately in each panel (value in brackets, log-odds), so each panel carries its own feature labels and the order differs between panels; the SHAP axis is common to all panels. Points are individual patients coloured by feature value (green = low, red = high; lavender = missing). The same analysis at AHI ≥15 and ≥30 is shown in Supplementary Figures S12–S15.

The BMI-stratified analysis (Figure 5e–h) showed a similar change in ranking. In the lowest BMI group (≤26 kg/m²), BMI ranked first by mean |SHAP|, but only because of a nearly constant negative contribution (about −0.7), and age showed the widest spread of SHAP values (Figure 5e). In the >26 to ≤30 and >30 to ≤34 kg/m² groups age ranked first, and in the latter BMI ranked fifth (Figure 5f, 5g). For participants with BMI >34 kg/m², BMI had the highest mean absolute SHAP value; its contributions were almost all positive, ranging from about 0 to +1.7 (Figure 5h), so the plots do not show a weaker BMI contribution in this group. Gender contributions were directionally consistent across BMI strata. The rankings at AHI ≥15 and ≥30 are shown in Supplementary Figures S12–S15; at AHI ≥30, BMI ranked first in the oldest age group and in the >34 kg/m² group.

SHAP dependence plots (Figure 6) showed nonlinear saturation effects for both age and BMI: SHAP contributions rose steeply across the lower ranges and flattened at higher values, turning positive at about 50 years of age and a BMI of about 32 kg/m². Interactions were modest in size (mean absolute SHAP interaction 0.113 for age × BMI, 0.104 for age × gender, 0.072 for BMI × gender and 0.041 for age × snoring, compared with mean |SHAP| of 0.527 for age and about 0.48 for BMI at AHI ≥5). Male sex shifted SHAP(BMI) upward over most of the observed BMI range without visibly changing the shape of the curve (Figure 6c). For age, by contrast, stratification by BMI or gender separated the curve vertically, with the ordering reversing at the oldest ages (above about 70 years; Figure 6a, 6b), indicating that the age contribution is modulated by both variables, whereas snoring level produced only a small vertical separation (Figure 6d). In the weaker pairs (Supplementary Figure S16), SHAP contributions of snoring increased with snoring intensity, with little change between levels 0 and 1 and a positive median contribution only from level 3 upward; the median contribution of witnessed apneas was positive from level 1 upward. High blood pressure, used as the stratified variable, did not visibly separate the age or BMI curves (Supplementary Figure S16e, S10f). These observations are descriptive and apply to the AHI ≥5 threshold only.

**Figure 6.**
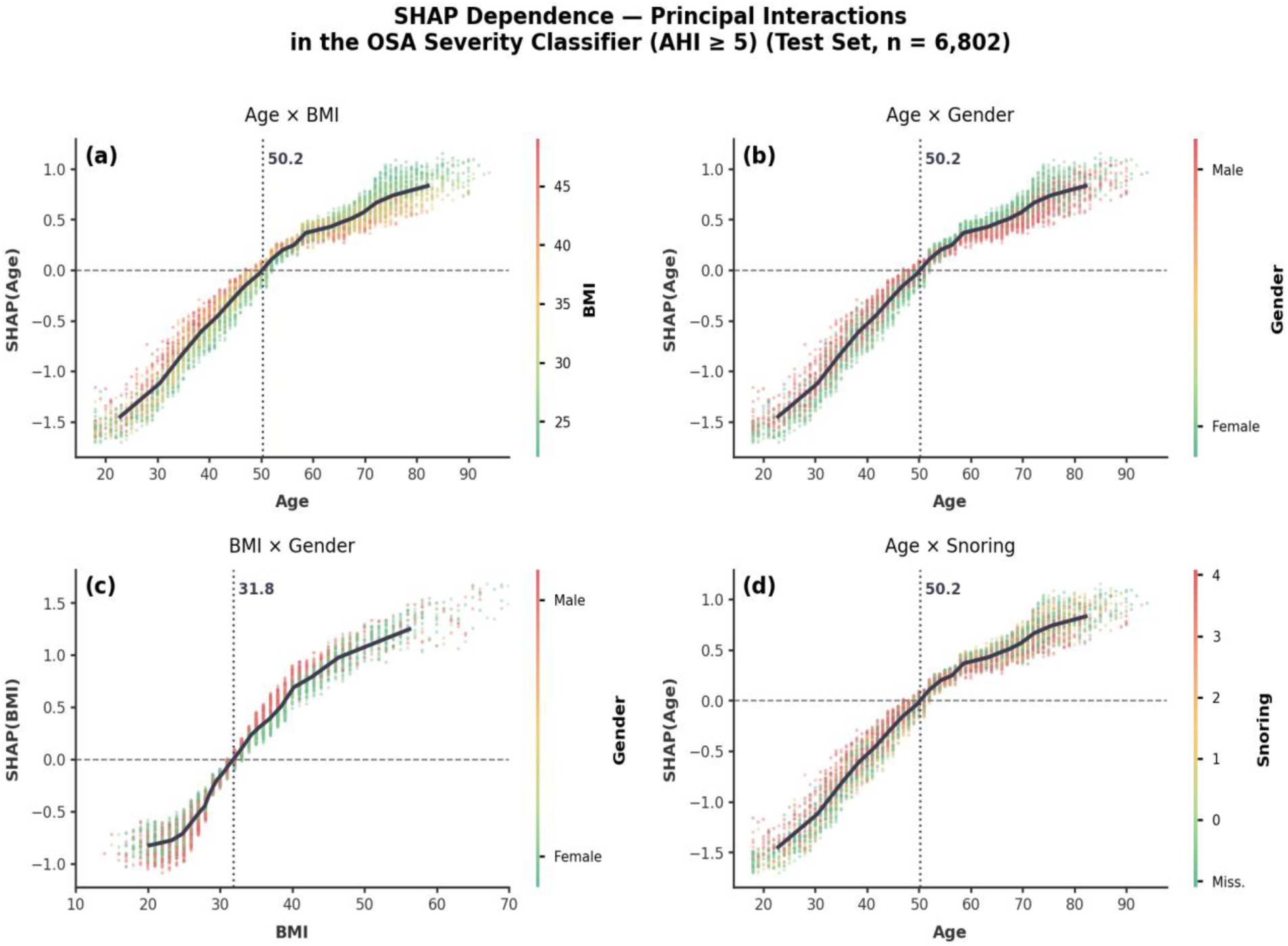
SHAP dependence plots for the four strongest pairwise interactions in the OSA severity classifier at AHI ≥5 (All-Features model, test set, n = 6,802). Panels: (a) Age × BMI, (b) Age × Gender, (c) BMI × Gender and (d) Age × Snoring; each panel plots the SHAP value of the first feature against its value, with points coloured by the second. Dashed horizontal line, SHAP = 0; solid line, mean SHAP in 20 equal-count bins; dotted vertical line, value at which mean SHAP becomes positive (age 50.2 years; BMI 31.8 kg/m²). Pairs were ranked by mean absolute SHAP interaction value on 1,000 randomly sampled test participants (log-odds: Age × BMI 0.113, Age × Gender 0.104, BMI × Gender 0.072, Age × Snoring 0.041; all other pairs ≤0.018); the next six pairs are shown in Supplementary Figure S16.

## 4. Discussion

This study provides a large within-cohort controlled comparison of full, questionnaire-only, and demographics-only feature sets for OSA severity classification to date. The four-variable demographic model (age, gender, BMI and race) had AUROCs 0.016–0.038 lower than the 169-feature model across all thresholds and indices. This difference was however small. The 95% confidence intervals excluded zero at four of the six endpoints, and the lower bound was approximately zero at AHI ≥5 and RDI ≥5. Further, the 161-variable questionnaire model excluding demographic variables performed consistently and substantially worse (see Supplementary Table S2 confidence intervals). Within this cohort and modelling framework, self-reported symptom data therefore added a small, measurable amount of discrimination once age, gender, BMI and race were available; whether a gain of this size is clinically useful is uncertain. These results should be interpreted in the context of the bounded reliability of PSG-derived labels: a deep-learning detector trained on full PSG signals reached R² = 0.84 agreement with expert-annotated AHI [26], which is consistent with label variability limiting the attainable performance of AHI-based classifiers.

Prior studies that included demographic variables alongside questionnaire items achieved AUROCs broadly comparable to the Demographics-Only model in the present cohort (Table 2). The present analysis, conducted within a single consistently defined and heterogenous cohort with explicit ablation of demographic and questionnaire feature sets, provides a more direct quantification of their respective contributions than is possible from cross-study comparison alone. The summary ROC DOR values of 4.8– 13.6 across thresholds, each based on three to six study cohorts, are consistent with the moderate overall discrimination observed here.

Three key gaps motivated the present study: (i) Üstün et al. [10] compared all-feature, medical-history/demographic and symptom-only models within one cohort; the present study extends that design to a larger cohort with explicit demographic and questionnaire ablations and a stratified SHAP analysis; (ii) existing cohorts are substantially smaller than the present sample; and (iii) the relative contribution of individual demographic predictors across patient subgroups has not been examined with stratified interpretability methods.

Respiratory events were scored according to standardized AASM criteria [25]. Apneas are defined as near-complete cessation of airflow lasting ≥10 seconds; hypopneas are defined as partial reductions in airflow accompanied by oxygen desaturation or arousal. Disease severity is summarized using the apnea–hypopnea index (AHI), which categorizes OSA as mild (5–14 events/h), moderate (15–29 events/h), or severe (≥30 events/h). The respiratory disturbance index (RDI) further incorporates respiratory effort-related arousals. While these metrics are central to clinical decision-making, AHI alone may not fully capture the heterogeneity of sleep-disordered breathing phenotypes or symptom burden [18]. Importantly, scoring of respiratory events is subject to substantial inter-rater variability [26], driven by subjectivity in baseline estimation, evolving hypopnea definitions, and site-specific practices. Scorer variability may limit attainable discrimination for any classifier trained on PSG-derived AHI; the size of this limit was not quantified here.

SHAP analysis confirmed demographic dominance but revealed an important subgroup pattern: the relative weighting of age versus BMI is not fixed but varies across age and BMI strata. This pattern is shown in Figure 5 and Supplementary Figures S12–S15. Whether this pattern generalizes beyond the present cohort requires validation in independent samples. The model associates female sex over 50 years with lower predicted OSA probability.

In the Questionnaire-Only model, high blood pressure ranked as the leading predictor, likely reflecting its epidemiological co-occurrence with OSA and partial proxying for the withheld demographic risk profile. Snoring and witnessed apneas were the only questionnaire items with consistent face validity as direct indicators of nocturnal respiratory disturbance. The remaining symptom features contributed diffuse, low-magnitude SHAP values without consistent directional patterns, indicating limited associations with objective AHI. Four-class classification yielded macro-averaged F1 scores ≤0.40 across all configurations, consistent with fine-grained severity stratification from non-PSG data being limited in this cohort (XGBoost accuracy did not exceed 0.45); binary threshold-based classification is therefore the more defensible clinical application.

Recent multimodal foundation models trained on full PSG signals (SleepFM [33]) achieve substantially higher discrimination—including accurate prediction of 130 conditions, all-cause mortality C-index 0.84, and sleep apnea severity classification accuracy of 0.69, which suggests that PSG signals carry more information than questionnaire and demographic data. The present findings identify the contribution from non-invasive, non-signal-based inputs and provide a reference point for the added value of full PSG acquisition.

These findings have direct implications for screening pathway design. Age, gender, and BMI are routinely recorded in clinical practice without requiring dedicated assessment, and the present results support prospective evaluation of demographic-first stratification approaches. Questionnaire administration retains clear value for symptom monitoring, treatment decision-making, and outcomes tracking. In contrast, a disease-specific questionnaire can be highly informative: in narcolepsy type 1, machine learning applied to a brief cataplexy questionnaire reached an AUROC of 0.995, against 0.863 for the Epworth Sleepiness Scale alone, with HLA-DQB1*06:02 typing mainly improving specificity [34]. This is consistent with questionnaires adding most when they capture a near-pathognomonic symptom, which the generic items available here may not. Two-stage pathways that follow a questionnaire with a low-burden objective measure have been proposed for isolated REM sleep behavior disorder [35]. For OSA, within the constraints of this analysis, adding extensive patient-reported symptom assessment to demographic variables improved pre-test discrimination of objective OSA severity only slightly (AUROC gain 0.016–0.038; approximate confidence intervals excluded zero at four of six endpoints). Whether a gain of this size justifies the burden of a long questionnaire is uncertain and should be tested prospectively, for example with decision-curve or cost-effectiveness analyses.

Six principal limitations should be considered when interpreting these findings. First, the retrospective design yields a referred population whose OSA prevalence and demographic composition may not reflect the general population, despite the multi-center nature of the dataset across US sleep centers operating under a standardized clinical protocol. Second, PSG-derived AHI varies between scorers because hypopnea identification is partly subjective and scoring definitions have changed [26]. Sites adopted successive AASM revisions during 2004–2019, so hypopnea criteria differed across the study period. These factors may limit classifier performance regardless of the feature set, but we did not quantify their effect. Third, AHI does not fully capture sleep-disordered breathing heterogeneity, including hypoxic burden, arousal frequency, and positional dependence; the predictability of these alternative metrics from questionnaire and demographic data was not examined. Fourth, SHAP rankings reflect statistical associations learned from training data within this specific modelling framework and do not establish causal relationships between predictors and OSA severity; demographic dominance in SHAP rankings should not be interpreted as evidence that questionnaire-derived symptoms are clinically uninformative in other contexts. Fifth, the questionnaire items reflect a custom legacy instrument designed for routine clinical use within this network and includes the eight Epworth Sleepiness Scale items and the total score but otherwise may not correspond precisely to validated instruments used in prior studies, such as the multidimensional Alliance Sleep Questionnaire [36], limiting direct comparisons of feature importance across settings. Notably, the questionnaire did not include the STOP-Bang or Berlin instruments or neck circumference, a predictor commonly reported in prior studies (Table 2), so our estimates may understate the value of validated screening tools. Sixth, predicted probabilities were not recalibrated: calibration slopes were close to 1 but intercepts were offset relative to class prevalence (Supplementary Table S9), so the decision-curve net-benefit estimates (Supplementary Tables S8–S9, Figure S8) remain provisional. Recalibration and prospective validation are needed before clinical use.

## 5. Conclusion

In 67,397 adults referred for diagnostic polysomnography, age, gender, BMI and race retained 86–94% of the above-chance AUROC (AUROC − 0.5) of a 169-feature model across all six endpoints, and adding the full questionnaire improved held-out AUROC by no more than 0.038 (Supplementary Table S2). Questionnaire-derived symptoms were therefore secondary to routinely recorded demographics for classifying OSA severity in this cohort, and no feature set predicted four-class severity well.

Practically, this suggests that questionnaire-based screening tools should be benchmarked against an age, gender and BMI baseline, and questionnaire length should be justified by incremental value. Stratified SHAP analyses showed that the contributions of age and BMI differ across strata in ways global rankings obscure, which may support stratum-specific screening thresholds. These conclusions come from a single referred cohort with a custom questionnaire and do not show that symptoms are clinically uninformative; prospective validation in independent, more diverse settings is the next step.

## CRediT authorship contribution statement

Rishabh Patted: Conceptualization, Data curation, Formal analysis, Investigation, Methodology, Visualization, Writing – original draft, Writing – review & editing.

Marcus Dige: Writing – original draft, Writing – review & editing.

Umaer Hanif: Writing – original draft, Writing – review & editing.

Emmanuel Mignot: Conceptualization, Funding acquisition, Resources, Supervision, Validation, Writing – original draft, Writing – review & editing, Resources.

Giorgio Ricciardiello Mejia: Conceptualization, Data curation, Formal analysis, Funding acquisition, Investigation, Methodology, Project administration, Resources, Supervision, Validation, Visualization, Writing – original draft, Writing – review & editing.

## Ethics statement

This retrospective study used intake-questionnaire and polysomnography records collected during routine clinical care. The study protocol was reviewed and approved by the Advarra institutional review board for the protection of human patients, as previously reported for this cohort [22], and the research was conducted in accordance with the ethical principles of the Declaration of Helsinki. All patients provided informed consent for the use of their data. The data were de-identified using the US Department of Health and Human Services (HIPAA) Safe Harbor method before analysis. The study was not a clinical trial and was not registered.

## Declaration of generative AI and AI-assisted technologies in the manuscript preparation process

During the preparation of this manuscript, the authors used ChatGPT GPT-6 (OpenAI) and Claude Sonnet 5.5 (Anthropic) to assist with language editing and improving the clarity of the text. All AI-assisted content was reviewed and revised by the authors, who take full responsibility for the final manuscript.

## Data availability statement

The de-identified analytical dataset underlying this study (subject-level demographic, questionnaire-derived and polysomnography-derived severity data; n = 67,397 adults) will be shared on PhysioNet [37] (Sleep Questionnaire and Demographic Data for Obstructive Sleep Apnea Research; DOI to be assigned at publication) upon publication, subject to a data use agreement. The dataset was de-identified following the HIPAA Safe Harbor method.

## Code availability statement

The analysis code used to produce the results reported here is available at https://github.com/GiorgioRicciardiello/QuestionnaireOsaSubjectiveBurden.

## References

[1] Benjafield AV, Ayas NT, Eastwood PR, Heinzer R, Ip MSM, Morrell MJ, et al. Estimation of the global prevalence and burden of obstructive sleep apnoea: a literature-based analysis. Lancet Respir Med 2019;7:687–98.

[2] Senaratna CV, Perret JL, Lodge CJ, Lowe AJ, Campbell BE, Matheson MC, et al. Prevalence of obstructive sleep apnea in the general population: A systematic review. Sleep Med Rev 2017;34:70–81. 10.1016/j.smrv.2016.07.002.

[3] Kendzerska T, Mollayeva T, Gershon AS, Leung RS, Hawker G, Tomlinson G. Untreated obstructive sleep apnea and the risk for serious long-term adverse outcomes: A systematic review. Sleep Med Rev 2014;18:49–59. 10.1016/j.smrv.2013.01.003.

[4] Ramar K, Malhotra RK, Carden KA, Martin JL, Abbasi-Feinberg F, Aurora RN, et al. Sleep is essential to health: an American Academy of Sleep Medicine position statement. J Clin Sleep Med 2021;17:2115–9.

[5] Suen C, Wong J, Ryan CM, Goh S, Got T, Chaudhry R, et al. Prevalence of undiagnosed obstructive sleep apnea among patients hospitalized for cardiovascular disease and associated in-hospital outcomes: a scoping review. J Clin Med 2020;9:989.

[6] Kapur VK, Auckley DH, Chowdhuri S, Kuhlmann DC, Mehra R, Ramar K, et al. Clinical practice guideline for diagnostic testing for adult obstructive sleep apnea: an American Academy of Sleep Medicine clinical practice guideline. J Clin Sleep Med 2017;13:479–504. 10.5664/jcsm.6506.

[7] Johns MW. A new method for measuring daytime sleepiness: the Epworth sleepiness scale. Sleep 1991;14:540–5.

[8] Netzer NC, Stoohs RA, Netzer CM, Clark K, Strohl KP. Using the Berlin Questionnaire to identify patients at risk for the sleep apnea syndrome. Ann Intern Med 1999;131:485–91.

[9] Chung F, Yegneswaran B, Liao P, Chung SA, Vairavanathan S, Islam S, et al. STOP questionnaire: a tool to screen patients for obstructive sleep apnea. Anesthesiology 2008;108:812–21.

[10] Ustun B, Westover MB, Rudin C, Bianchi MT. Clinical prediction models for sleep apnea: the importance of medical history over symptoms. J Clin Sleep Med 2016;12:161–8.

[11] Wu MF, Huang WC, Juang CF, Chang KM, Wen CY, Chen YH, et al. A new method for self-estimation of the severity of obstructive sleep apnea using easily available measurements and neural fuzzy evaluation system. IEEE J Biomed Health Inform 2017;21:1524–32.

[12] Huang WC, Lee PL, Liu YT, Chiang AA, Lai F. Support vector machine prediction of obstructive sleep apnea in a large-scale Chinese clinical sample. Sleep 2020;43:zsz295.

[13] Kim YJ, Jeon JS, Cho SE, Kim KG, Kang SG. Prediction models for obstructive sleep apnea in korean adults using machine learning techniques. Diagnostics 2021;11:612. 10.3390/diagnostics11040612.

[14] Ramesh J, Keeran N, Sagahyroon A, Aloul F. Towards validating the effectiveness of obstructive sleep apnea classification from electronic health records using machine learning. Healthcare 2021;9:1450. 10.3390/healthcare9111450.

[15] Tsai CY, Liu WT, Lin YT, Lin SY, Houghton R, Hsu WH, et al. Machine learning approaches for screening the risk of obstructive sleep apnea in the Taiwan population based on body profile. Inform Health Soc Care 2022;47:373–88.

[16] Holfinger SJ, Lyons MM, Keenan BT, Mazzotti DR, Mindel J, Maislin G, et al. Diagnostic Performance of Machine Learning-Derived OSA Prediction Tools in Large Clinical and Community-Based Samples. Chest 2022;161:807–17. 10.1016/j.chest.2021.10.023.

[17] Han H, Oh J. Application of various machine learning techniques to predict obstructive sleep apnea syndrome severity. Sci Rep 2023;13:6379.

[18] Huo J, Quan SF, Roveda J, Li A. BASH-GN: a new machine learning–derived questionnaire for screening obstructive sleep apnea. Sleep Breath 2023;27:449–57.

[19] Maniaci A, Riela PM, Iannella G, Lechien JR, La Mantia I, De Vincentiis M, et al. Machine learning identification of obstructive sleep apnea severity through the patient clinical features: a retrospective study. Life (Basel) 2023;13:702.

[20] Liu K, Geng S, Shen P, Zhao L, Zhou P, Liu W. Development and application of a machine learning-based predictive model for obstructive sleep apnea screening. Front Big Data 2024;7:1353469.

[21] Russo S, Martini A, Luzzi V, Garbarino S, Pietrafesa E, Polimeni A. Exploring the complexity of obstructive sleep apnea: findings from machine learning on diagnosis and predictive capacity of individual factors. Sleep Breath 2025;29:49.

[22] Hanif U, Cairns A, Mysliwiec V, Bettinardi RG, Elbaz M, Gimenez U, et al. Associations between self-reported parasomnias and psychiatric illness in 370,000 patients with sleep disorders. Psychiatry Clin Neurosci 2024;78:667–77.

[23] Page MJ, McKenzie JE, Bossuyt PM, Boutron I, Hoffmann TC, Mulrow CD, et al. Updating guidance for reporting systematic reviews: development of the PRISMA 2020 statement. J Clin Epidemiol 2021;134:103–12.

[24] Sayers A. Tips and tricks in performing a systematic review. Br J Gen Pract 2008;58:136.

[25] Berry RB, Budhiraja R, Gottlieb DJ, Gozal D, Iber C, Kapur VK, et al. Rules for scoring respiratory events in sleep: update of the 2007 AASM Manual for the Scoring of Sleep and Associated Events. Deliberations of the Sleep Apnea Definitions Task Force of the American Academy of Sleep Medicine. J Clin Sleep Med 2012;8:597–619.

[26] Kjaer MR, Hanif U, Brink-Kjaer A, Olsen M, Sum-Ping O, Carrillo O, et al. Expert-level probabilistic breathing event detector informs phenotyping of sleep apnea. Nat Commun 2026;17. 10.1038/s41467-026-69163-z.

[27] Berry RB, Brooks R, Gamaldo CE, et al. The AASM Manual for the Scoring of Sleep and Associated Events: Rules, Terminology and Technical Specifications. Version 2.4. Darien, IL: American Academy of Sleep Medicine; 2017.

[28] Ricciardiello G, Patted R. QuestionnaireOsaSubjectiveBurden. GitHub 2026. https://github.com/GiorgioRicciardiello/QuestionnaireOsaSubjectiveBurden.

[29] Chen T, Guestrin C. XGBoost: A Scalable Tree Boosting System. Proceedings of the 22nd ACM SIGKDD International Conference on Knowledge Discovery and Data Mining. New York, NY, USA: ACM; 2016, p. 785–94. 10.1145/2939672.2939785.

[30] Akiba T, Sano S, Yanase T, Ohta T, Koyama M. Optuna: A Next-generation Hyperparameter Optimization Framework. Proceedings of the 25th ACM SIGKDD International Conference on Knowledge Discovery and Data Mining. New York, NY, USA: ACM; 2019, p. 2623–31. 10.1145/3292500.3330701.

[31] Hollmann N, Müller S, Eggensperger K, Hutter F. TabPFN: A transformer that solves small tabular classification problems in a second. International Conference on Learning Representations (ICLR); 2023. https://arxiv.org/abs/2207.01848.

[32] Prior Labs Team. TabPFN-2.5: Advancing the State of the Art in Tabular Foundation Models. arXiv:2511.08667; 2025. https://arxiv.org/abs/2511.08667.

[33] Thapa R, Kjaer MR, He B, Covert I, Moore IV H, Hanif U, et al. A multimodal sleep foundation model for disease prediction. Nat Med 2026;32:752–62. 10.1038/s41591-025-04133-4.

[34] Mejia GR, Brink-Kjaer A, Mignot E. Classification of narcolepsy type 1 using machine learning on Stanford Cataplexy Questionnaire responses and HLA-DQB1*06:02. Sleep Med 2026;143:108927. 10.1016/j.sleep.2026.108927.

[35] Massimi CA, Mejia GR, Metzger A, Ryu KH, Marwaha S, Grzegorczyk E, et al. A two-stage questionnaire and actigraphy screening for iRBD in a multicenter retrospective cohort. Ann Clin Transl Neurol 2026. 10.1002/acn3.70439.

[36] Wilson A, Ricciardiello Mejia GC, Lomba S, Geng LN, Malunjkar S, Bonilla H, et al. A multidimensional assessment of sleep disorders in long COVID using the Alliance Sleep Questionnaire. Healthcare 2025;13:2611. 10.3390/healthcare13202611.

[37] Dige M, et al. Sleep Questionnaire and Demographic Data for Obstructive Sleep Apnea Research. PhysioNet; forthcoming (DOI to be assigned at publication).

