## Supplementary Material for "Do Sleep Questionnaires Add Predictive Value Beyond Demographics for Obstructive Sleep Apnea Severity? Evidence from 67,397 Patients"

### Supplementary Materials

This document contains supplementary figures and tables referenced in the main manuscript. Figure S1 and Table S1 trace the derivation of the analytic cohort from the source records. Figures S5–S7 relate to the systematic review and meta-analysis; Tables S7, S10, and S12 report complete model performance metrics; Tables S2–S3 and S13 report approximate confidence intervals, majority-class baselines and SHAP concentration summaries computed post hoc; remaining figures and tables report SHAP-based interpretability analyses and data quality checks.

#### Supplementary Figures

##### Figure S1. Cohort Data Flow

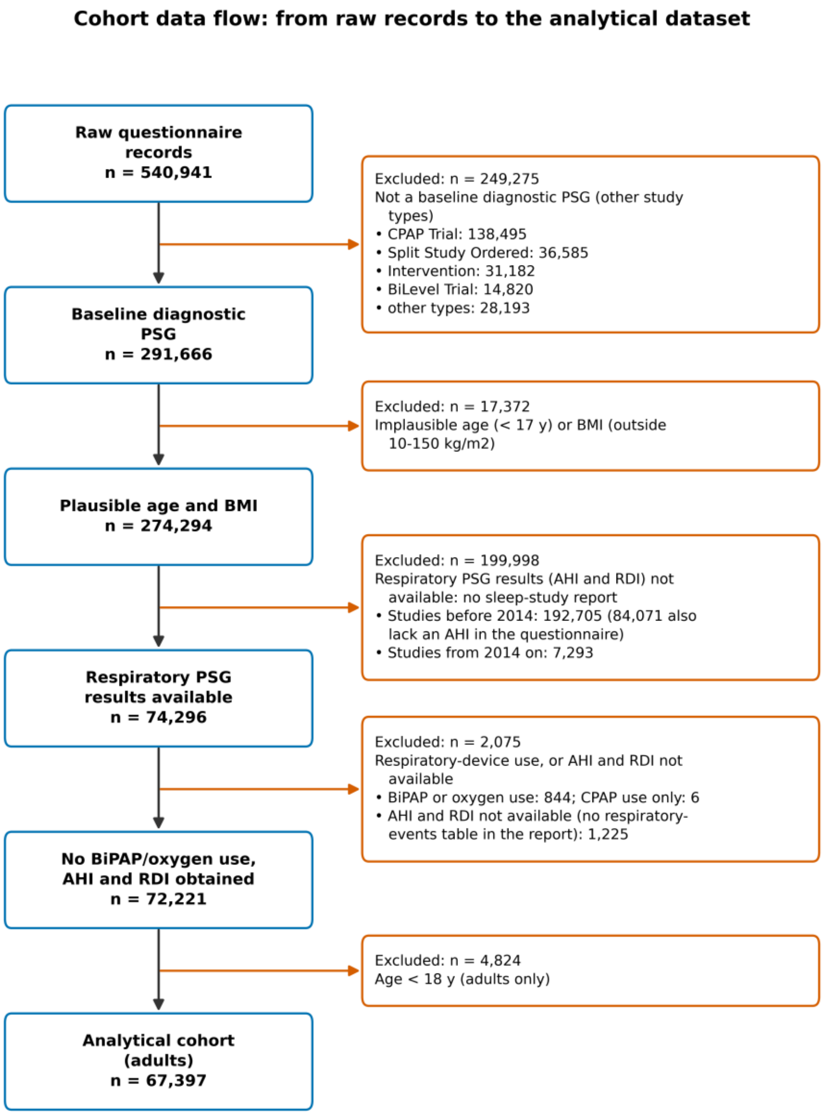


Flow of records from the source questionnaire database (540,941 records) to the analytical cohort (67,397 adults). Blue boxes give the records remaining after each step; orange boxes give the number excluded and the rule applied. AHI and RDI came from sleep-study reports, available for studies from 2014 onwards. Step 4 excluded BiPAP or oxygen users (and six CPAP-only users) and reports without a respiratory-events table; the remaining CPAP-only users were retained. No patient was excluded for missing questionnaire items. AHI, apnea–hypopnea index; BiPAP, bilevel positive airway pressure; BMI, body mass index; CPAP, continuous positive airway pressure; PSG, polysomnography; RDI, respiratory disturbance index.

#### Figure S2. Stratification Variable Distributions — Binary Target (RDI ≥5), First Split

*Distribution of stratification variables across full, training (90%), and test (10%) splits for the binary RDI ≥5 target in the first 90/10 development/test split. Panels: (a) age, (b) gender, (c) BMI, (d) race, (e) snoring score, (f) witnessed apneas, (g) lab site, (h) physician. Consistent distributions across splits (maximum Δ ≤0.49 percentage points) confirm that stratification preserved demographic and clinical feature balance.*

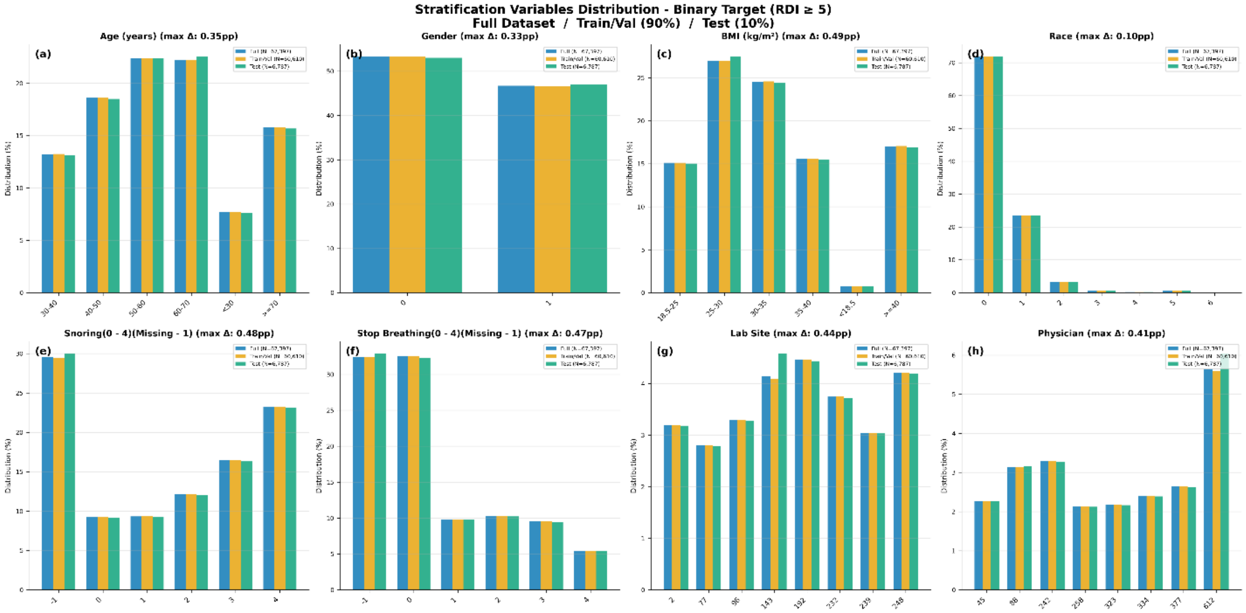


#### Figure S3. Stratification Variable Distributions — Multiclass Target (RDI 4-Class), First Split

*Distribution of stratification variables across the full dataset, training (90%), and test (10%) splits for the multiclass RDI target in the first 90/10 development/test split. Panels: (a) age, (b) gender, (c) BMI, (d) race, (e) snoring score, (f) witnessed apneas, (g) lab site, (h) referring physician. Distributions are highly consistent across splits (maximum Δ ≤0.72 percentage points), confirming effective stratified sampling.*

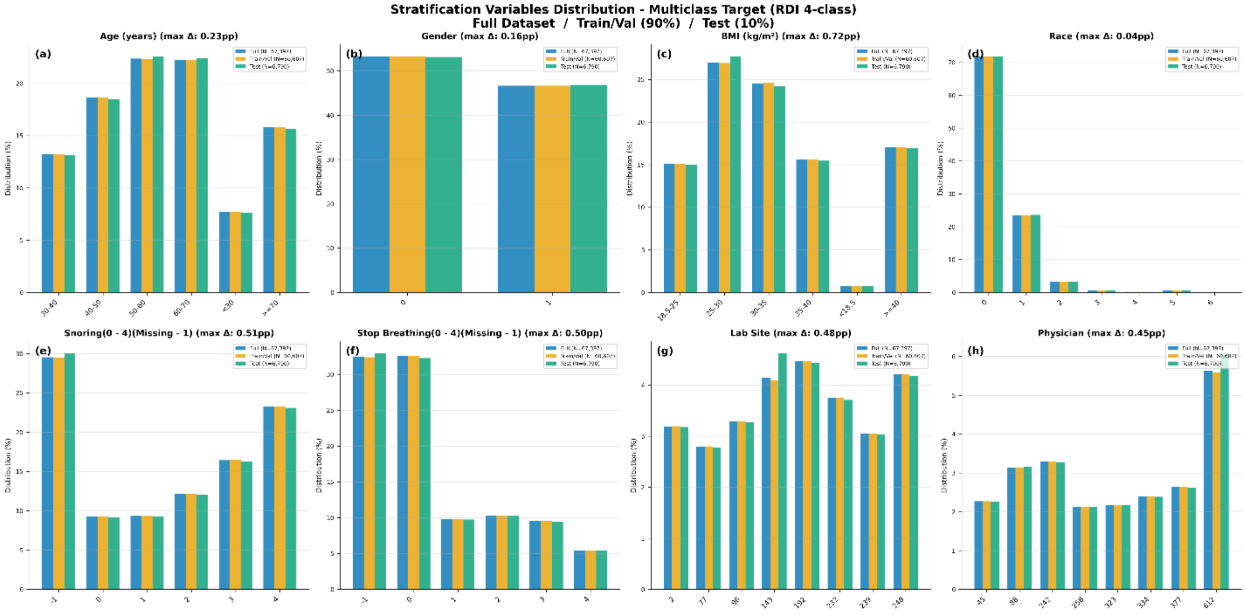

#### Figure S4. Target Class Distribution — Binary (RDI ≥5) and Multiclass Target, First Split

*Target class distribution across full, training, and test splits for the first 90/10 development/test split. (a) Binary classification target (RDI ≥5); (b) multiclass severity distribution (None, Mild, Moderate, Severe). Class proportions are well preserved across splits (maximum Δ ≤0.59 percentage points), confirming representative distributions in training and evaluation sets.*

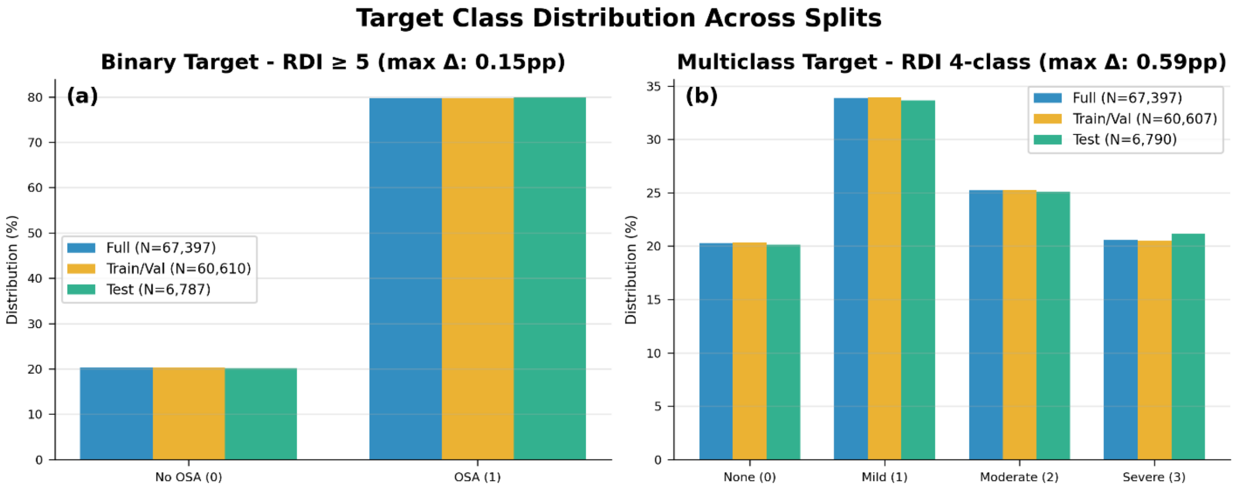


#### Figure S5. PRISMA Flow Diagram

*Records were identified from three databases (Google Scholar, PubMed, and Embase), screened for relevance, and filtered based on predefined criteria. Inclusion criteria: (i) ML models for OSA classification based on sleep questionnaire data; (ii) in-laboratory PSG as the reference standard; (iii) peer-reviewed publication; (iv) adult population (age ≥18 years). Exclusion criteria: (i) studies using physiological signals (ECG, $SpO_2$); (ii) non-ML statistical models. After full-text assessment, 12 studies were included, categorised by AHI cut-offs.*

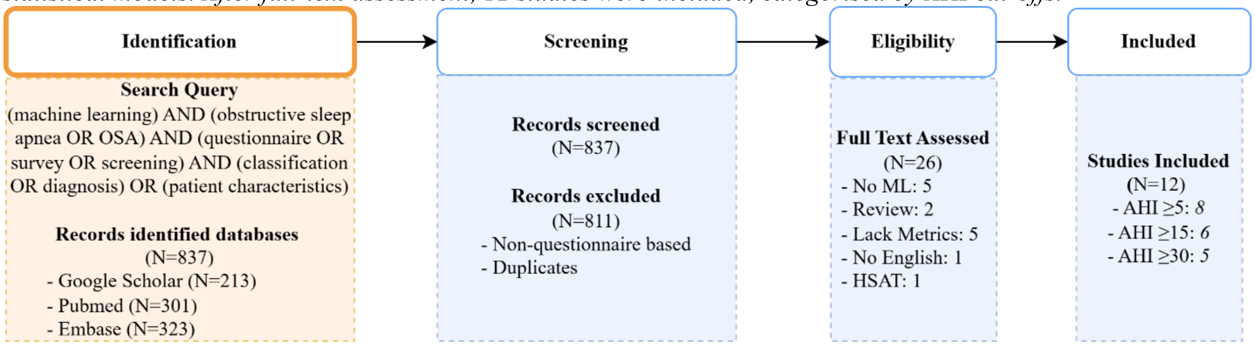

#### Figure S6. Summary ROC Curves

*Summary receiver operating characteristic (sROC) curves for ML-based screening models at AHI thresholds ≥5, ≥15 and ≥30 events/h. Each point is one study cohort, plotted by false positive rate (1 − specificity) and true positive rate (sensitivity), using the values and thresholds in manuscript Table 2 (AHI ≥5: k = 4; AHI ≥15: k = 6; AHI ≥30: k = 3). A study is plotted only at the threshold used in the source publication and only when both sensitivity and specificity were reported, so Kim et al., Han & Oh and Liu et al. are not shown; Wu et al. and both Huo et al. cohorts are plotted at AHI ≥15, as in the source studies. sROC curves were estimated with the unweighted Moses–Littenberg model, in which the diagnostic log odds ratio (D) is regressed on the accuracy–threshold parameter (S); parameters are given in Table S6. *Üstün et al. defined OSA as AHI >5 or RDI >10. †Maniaci et al. classified severe (AHI ≥30) versus mild-to-moderate OSA, so their sample excludes patients without OSA; the reduced logistic-regression model is shown. With three to six cohorts per threshold the curves are imprecise and should be compared descriptively.*

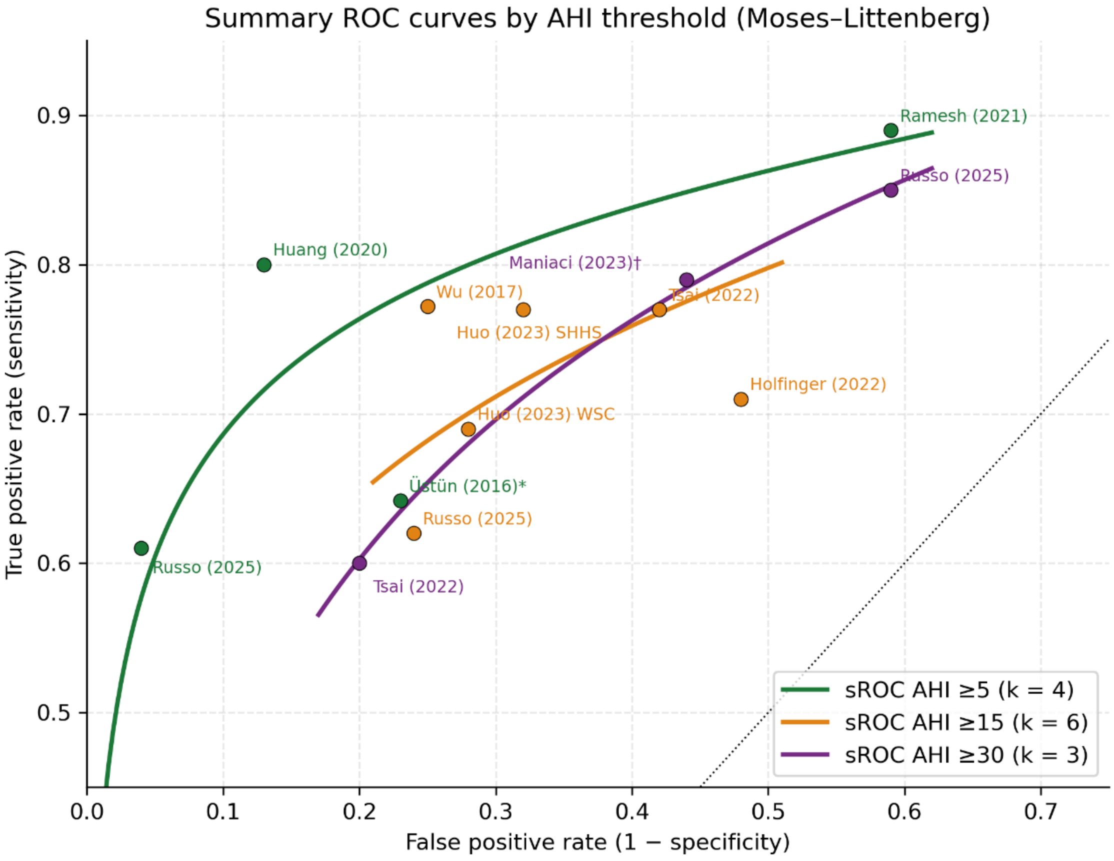

#### Figure S7. Bivariate Sensitivity–Specificity Summary

*Bivariate summary of sensitivity and specificity for ML-based screening models at AHI thresholds ≥5, ≥15 and ≥30 events/h, using the study cohorts in Figure S6. Small points are individual cohorts; large points are summary estimates, computed as the mean logit sensitivity and mean logit false positive rate across cohorts and back-transformed (AHI ≥5: sensitivity 0.76, specificity 0.81; AHI ≥15: 0.73, 0.67; AHI ≥30: 0.76, 0.60). Each threshold has a 95% confidence ellipse for its summary point, based on the between-study covariance of the logits; within-study sampling error was not modelled because 2×2 counts were not available for all studies, so this is a simplified alternative to a bivariate random-effects (Reitsma) model. AHI ≥5 shows the highest summary specificity and the highest diagnostic odds ratio, both at the mean S and at the Q-point (Table S6). The ellipses are wide and overlap, reflecting the small number of cohorts per threshold, so differences between thresholds are not established.*

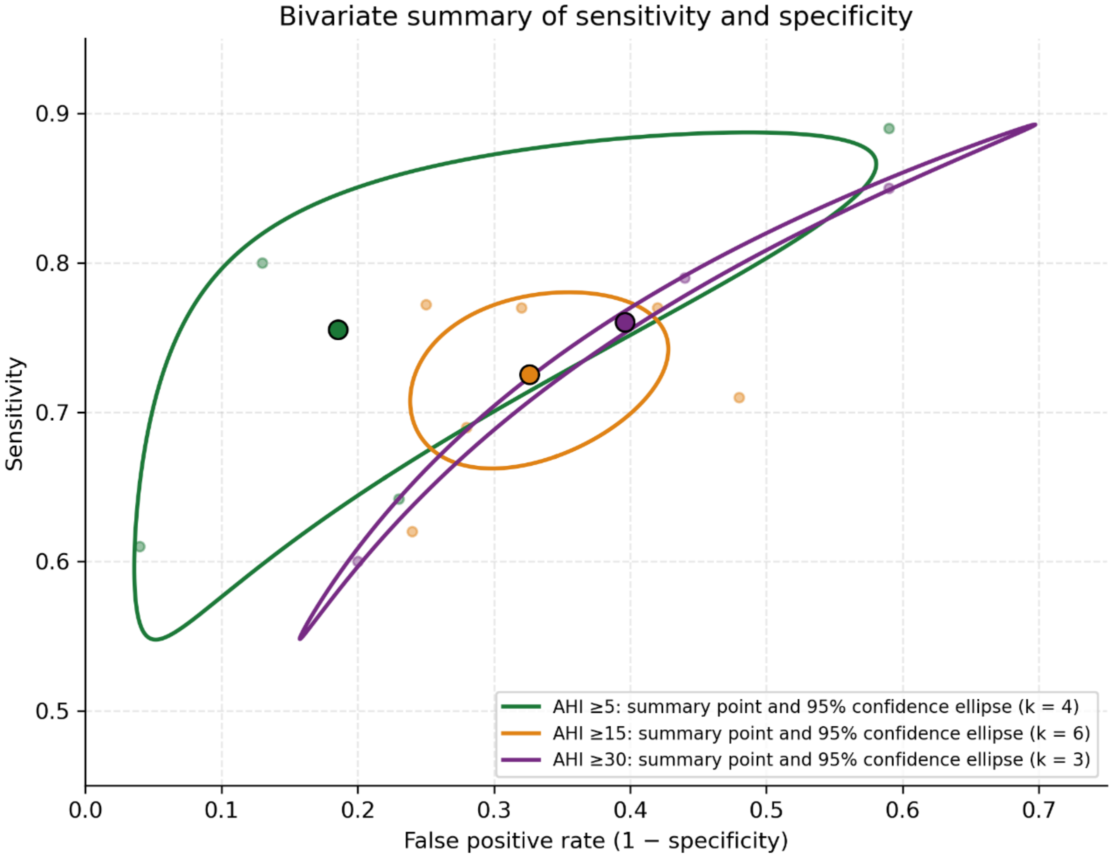

#### Figure S8. Decision curves and paired net-benefit differences

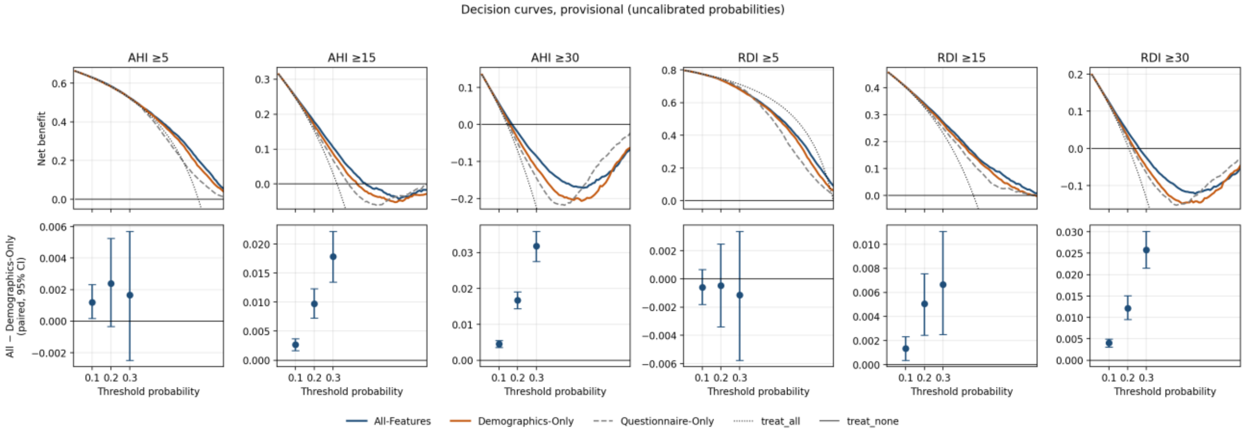


Top row: net benefit against threshold probability for All-Features, Demographics-Only and Questionnaire-Only XGBoost models, with treat-all (dotted) and treat-none (solid) references, on the held-out test set of each endpoint. Curves are clipped at the lower edge of each panel. Bottom row: paired difference in net benefit, All-Features minus Demographics-Only, at thresholds 0.10, 0.20 and 0.30 with 95% percentile intervals (Supplementary Table S8). Probabilities are uncalibrated; see Supplementary Methods and Supplementary Table S9.

#### Figure S9. XGBoost Four-Class Classification Confusion Matrices

*Confusion matrices of the XGBoost All-Features classifier for four-class sleep apnea severity prediction (None, Mild, Moderate, Severe) on the held-out test set. (a) AHI-based classification (n = 6,775); (b) RDI-based classification (n = 6,790). Rows represent true severity class; columns represent predicted class. Each cell reports absolute count and proportion within the true class. Predictions are assigned to the class with the highest predicted probability (argmax). These matrices give the All-Features values in Table S10 (AHI: accuracy 0.446, macro-F1 0.378; RDI: accuracy 0.417, macro-F1 0.399). Correct classification concentrates in the None and Mild classes for AHI (recall 0.59 and 0.55) and in the Mild and Severe classes for RDI (recall 0.61 and 0.44). Moderate disease was rarely identified (recall 0.09 for AHI and 0.20 for RDI), and most moderate cases, and many severe cases, were predicted as Mild.*

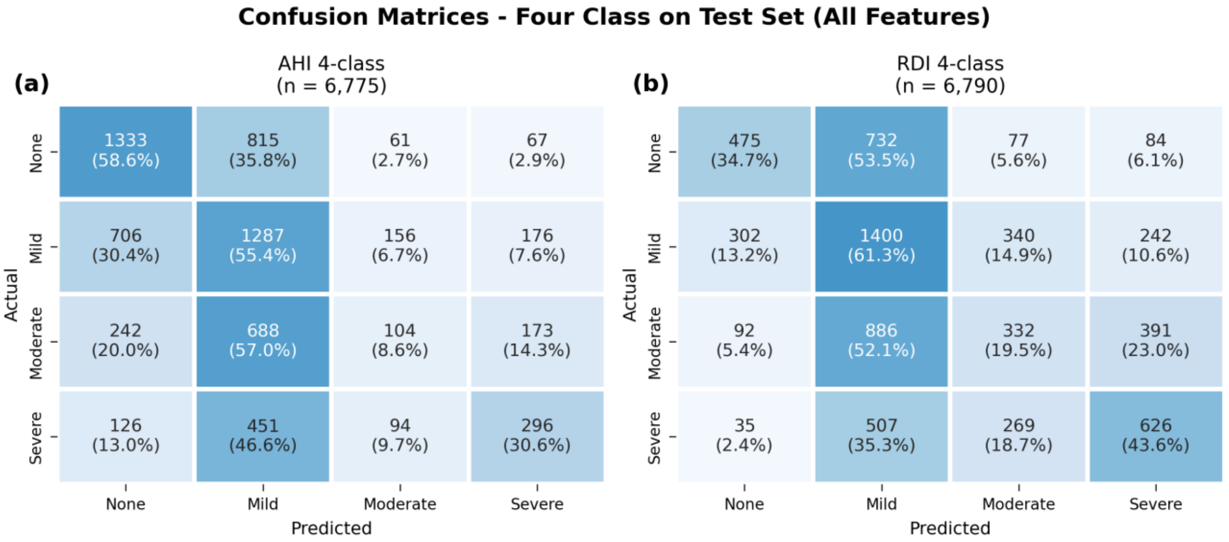

#### Figure S10. SHAP Feature Importance — Four-Class AHI Model (All Features)

*SHAP beeswarm plots showing the distribution of feature contributions to the four-class AHI severity model (None, Mild, Moderate, Severe) trained using the All-Features configuration, in which demographic and questionnaire-derived variables were all obtained from the same patient questionnaire. Each panel corresponds to one predicted class, with its AHI cut-offs given in the panel title: (a) No OSA, (b) Mild OSA, (c) Moderate OSA, (d) Severe OSA. Values in brackets are the mean absolute SHAP value (log-odds) of each feature for that class; the SHAP axes differ between panels. Features are ranked by mean absolute SHAP value. Age and BMI rank among the most influential predictors in every class; gender ranks among the top three for No OSA, Moderate OSA and Severe OSA but falls outside the top ten for Mild OSA, where difficulty falling asleep is the leading predictor. Questionnaire-reported symptoms including snoring, witnessed apneas, sleep difficulty, and hypertension make secondary contributions overall. Abbreviations: SHAP, SHapley Additive exPlanations; AHI, apnea–hypopnea index.*

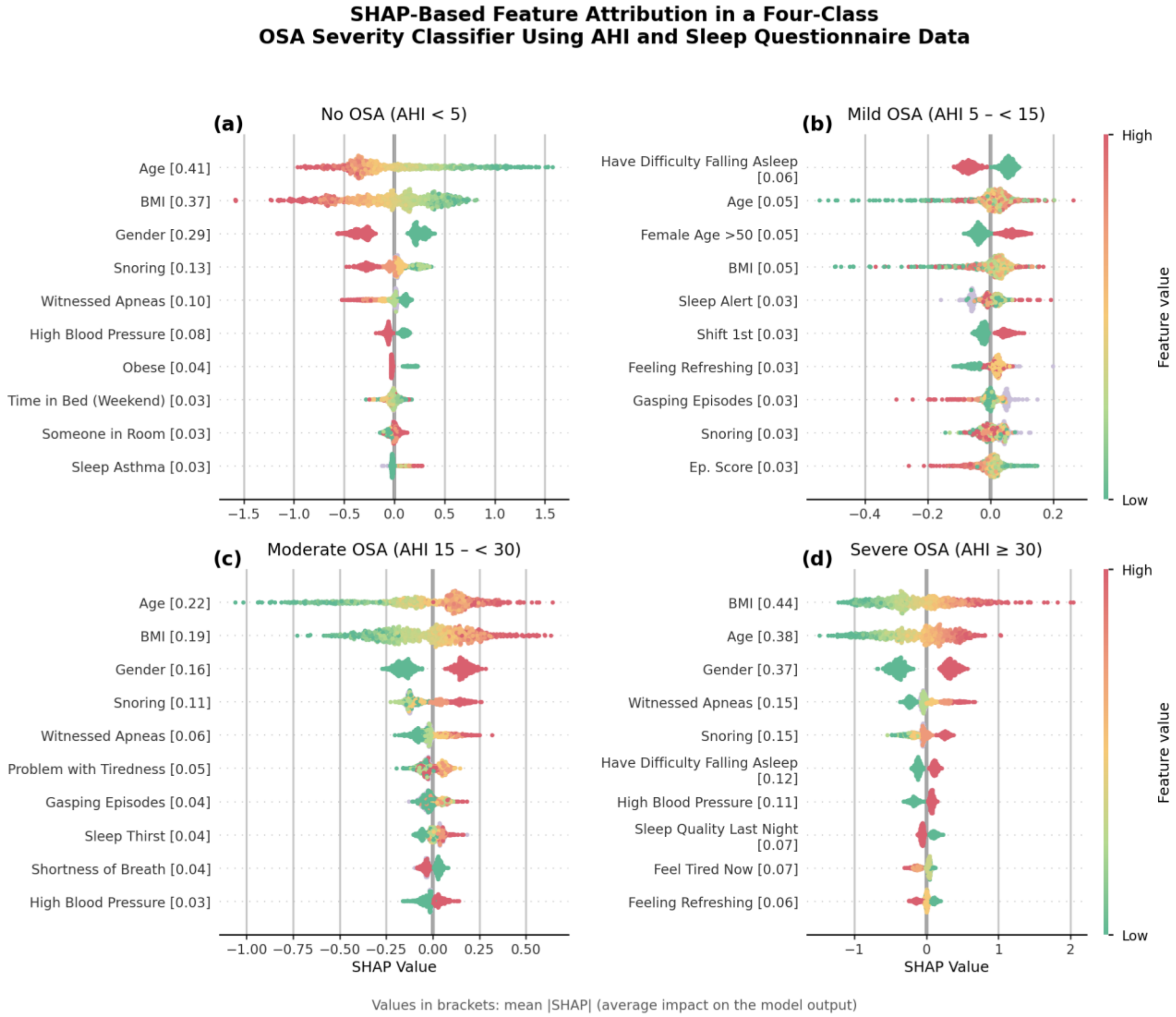


#### Figure S11. SHAP Feature Importance — Four-Class RDI Model (All Features)

*SHAP beeswarm plots showing the distribution of feature contributions to the four-class RDI severity model (None, Mild, Moderate, Severe) trained using the All-Features configuration, in which demographic and questionnaire-derived variables were all obtained from the same patient questionnaire. Each panel corresponds to one predicted class, with its RDI cut-offs given in the panel title: (a) No OSA, (b) Mild OSA, (c) Moderate OSA, (d) Severe OSA. Values in brackets are the mean absolute SHAP value (log-odds) of each feature for that class; the SHAP axes differ between panels. Features are ranked by mean absolute SHAP value. Age, BMI, and gender consistently rank among the most influential predictors; questionnaire-reported symptoms provide secondary contributions. Abbreviations: SHAP, SHapley Additive exPlanations; RDI, respiratory disturbance index.*

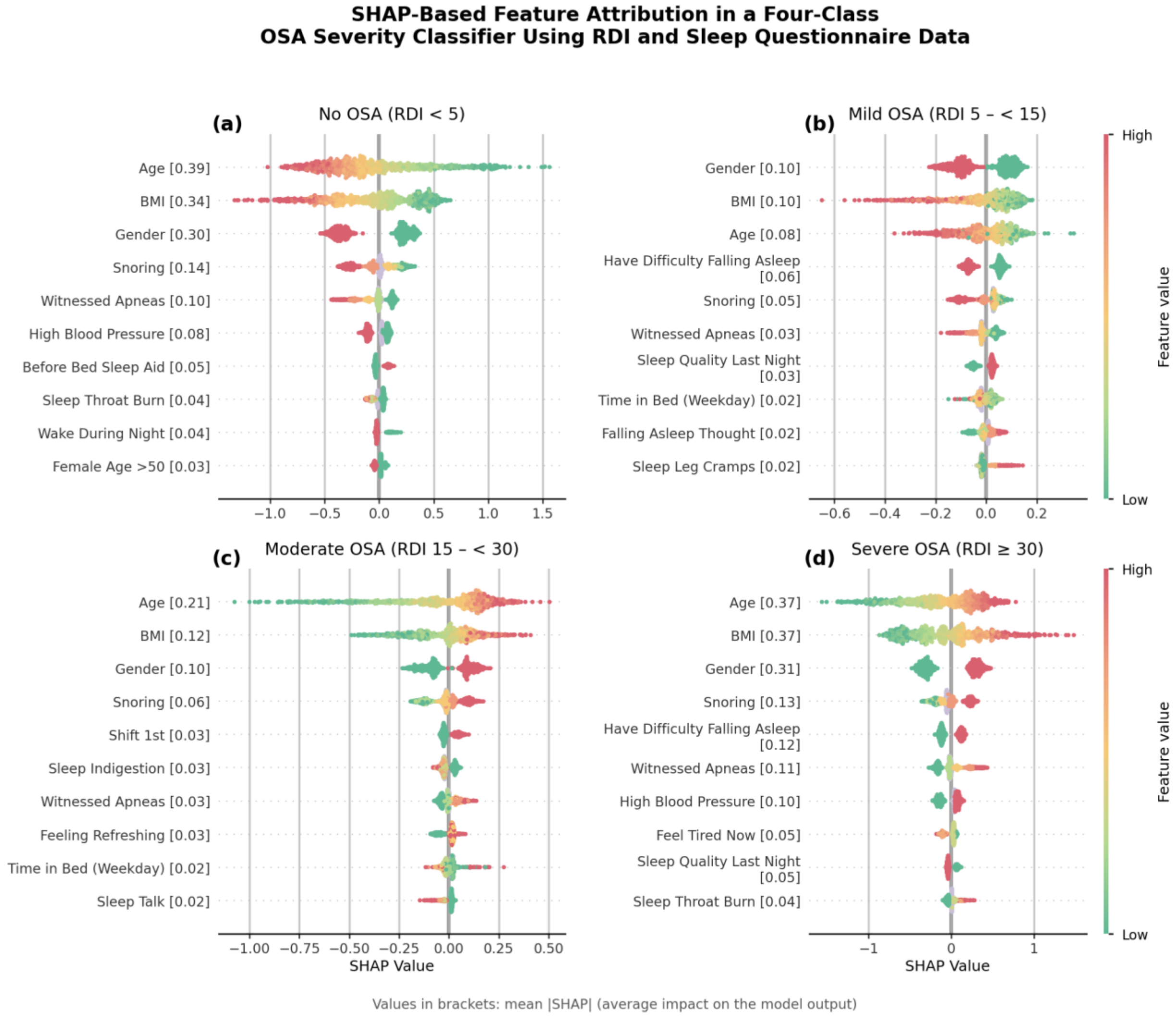

#### Figure S12. SHAP Beeswarm by Age Group — AHI ≥15

*SHAP summary plots for the All-Features XGBoost binary model at AHI ≥15 (held-out test set), stratified by age group (≤ 36, > 36 to ≤ 46, > 46 to ≤ 58, > 58 years). Panels (a)–(d) show the four strata and n is the number of test-set patients in the stratum. Features were ranked by mean absolute SHAP value separately in each panel (value in brackets, log-odds), so each panel carries its own feature labels. Points are individual patients coloured by feature value (green = low, red = high; lavender = missing). The AHI ≥5 results are shown in main-text Figure 5.*

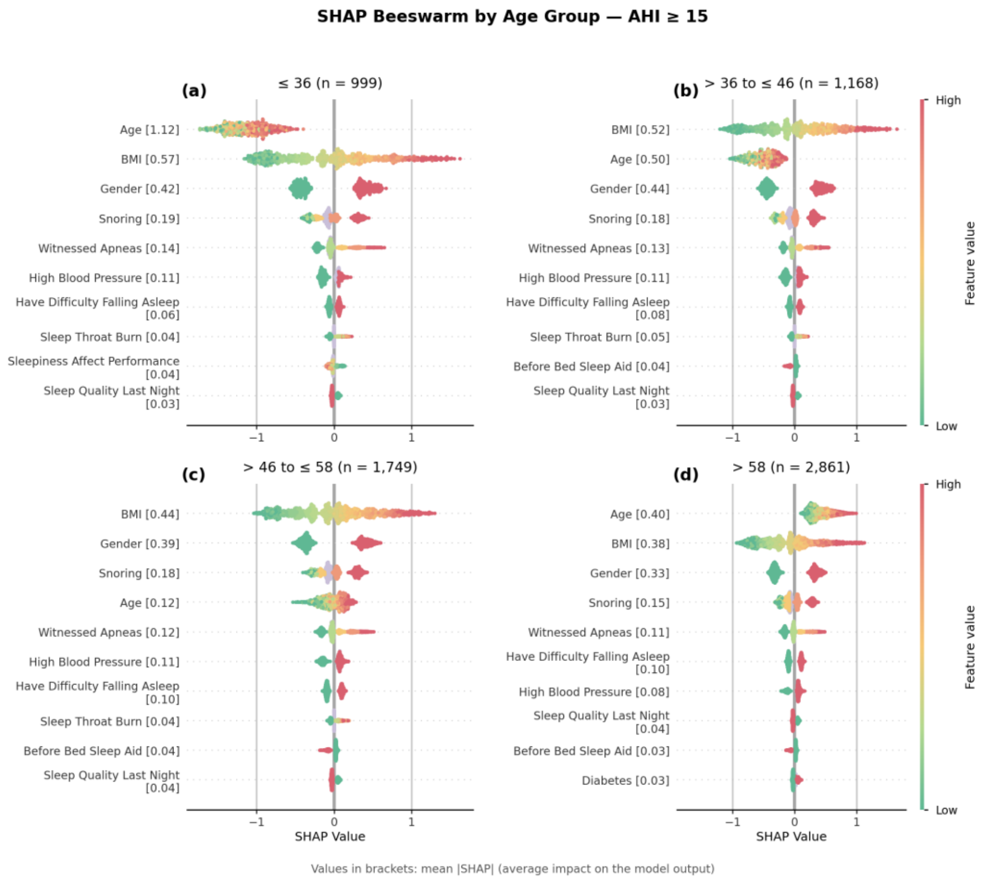


#### Figure S13. SHAP Beeswarm by Age Group — AHI ≥30

*SHAP summary plots for the All-Features XGBoost binary model at AHI ≥30 (held-out test set), stratified by age group (≤ 36, > 36 to ≤ 46, > 46 to ≤ 58, > 58 years). Panels (a)–(d) show the four strata and n is the number of test-set patients in the stratum. Features were ranked by mean absolute SHAP value separately in each panel (value in brackets, log-odds), so each panel carries its own feature labels. Points are individual patients coloured by feature value (green = low, red = high; lavender = missing). The AHI ≥5 results are shown in main-text Figure 5.*

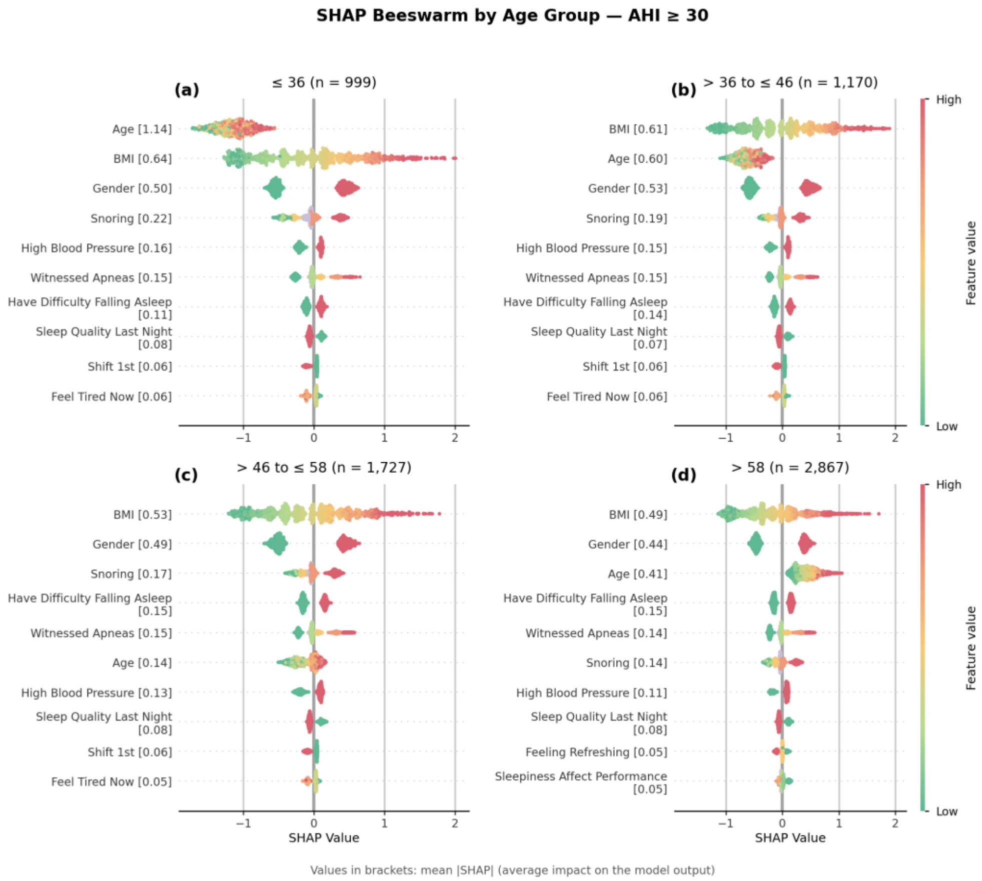


#### Figure S14. SHAP Beeswarm by BMI Group — AHI ≥15

*SHAP summary plots for the All-Features XGBoost binary model at AHI ≥15 (held-out test set), stratified by BMI category (≤ 26, > 26 to ≤ 30, > 30 to ≤ 34, > 34 kg/m²). Panels (a)–(d) show the four strata and n is the number of test-set patients in the stratum. Features were ranked by mean absolute SHAP value separately in each panel (value in brackets, log-odds), so each panel carries its own feature labels. Points are individual patients coloured by feature value (green = low, red = high; lavender = missing). The AHI ≥5 results are shown in main-text Figure 5.*

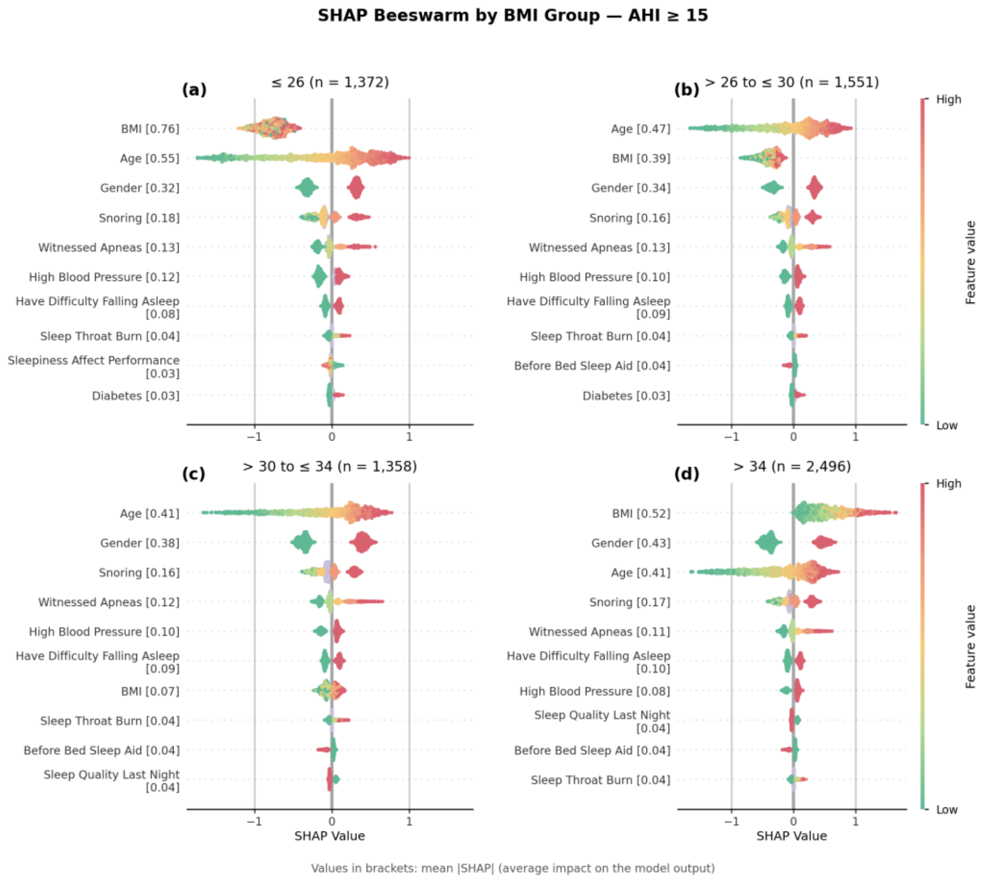


#### Figure S15. SHAP Beeswarm by BMI Group — AHI ≥30

*SHAP summary plots for the All-Features XGBoost binary model at AHI ≥30 (held-out test set), stratified by BMI category (≤ 26, > 26 to ≤ 30, > 30 to ≤ 34, > 34 kg/m²). Panels (a)–(d) show the four strata and n is the number of test-set patients in the stratum. Features were ranked by mean absolute SHAP value separately in each panel (value in brackets, log-odds), so each panel carries its own feature labels. Points are individual patients coloured by feature value (green = low, red = high; lavender = missing). The AHI ≥5 results are shown in main-text Figure 5.*

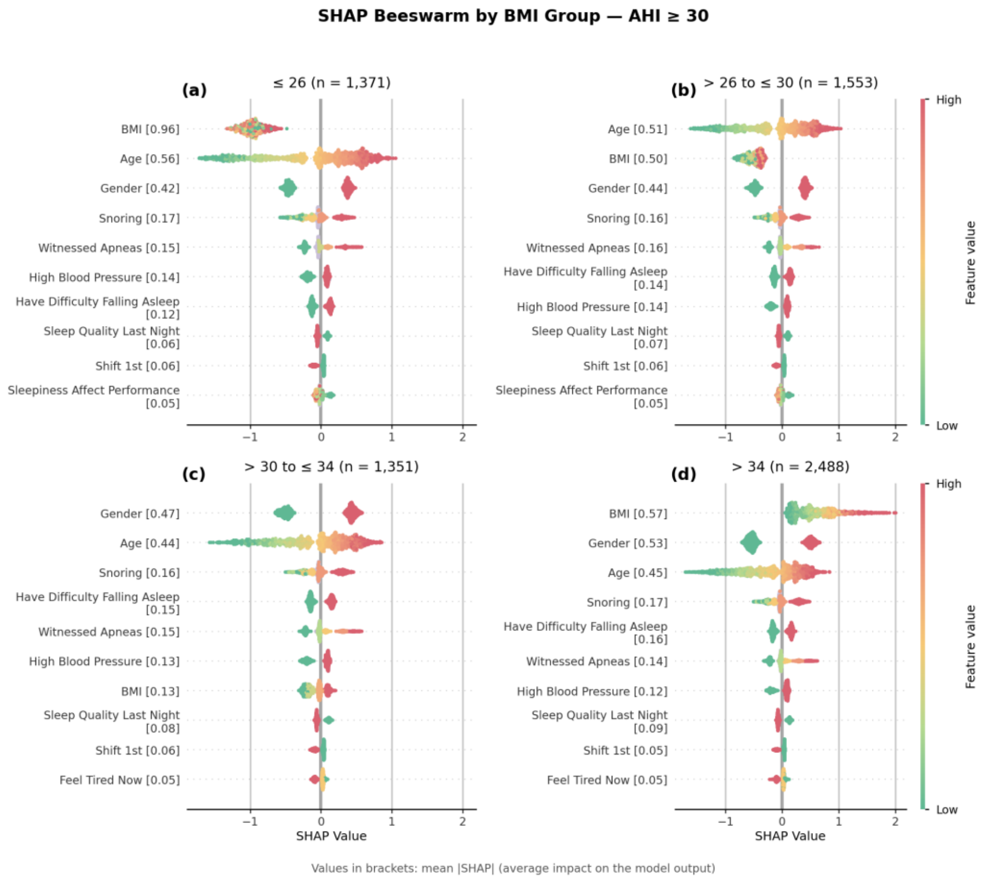


#### Figure S16. SHAP Dependence — Additional Pairwise Interactions (AHI ≥5)

*SHAP dependence plots for the weaker pairwise interactions in the AHI ≥5 XGBoost All-Features model (held-out test set, n = 6,802; mean absolute SHAP interaction 0.011–0.018 log-odds). Panels: (a) Snoring × BMI, (b) Snoring × Witnessed Apneas, (c) Snoring × Gender, (d) Witnessed Apneas × BMI, (e) Age × High Blood Pressure, (f) BMI × High Blood Pressure. Each panel plots the SHAP value of the first feature against its value, with points coloured by the second; discrete variables are jittered horizontally and "Missing" denotes unanswered items. Dashed horizontal line, SHAP = 0; solid line, mean SHAP in 20 equal-count bins; horizontal bars, median SHAP per category with n; dotted vertical line, value at which the mean (or category median) SHAP becomes positive. Interaction strengths were computed on 1,000 randomly sampled test participants. The four strongest interactions are shown in main-text Figure 6.*

**SHAP Dependence — Additional Interactions in the OSA Severity Classifier (AHI ≥ 5) (Test Set, n = 6,802)**

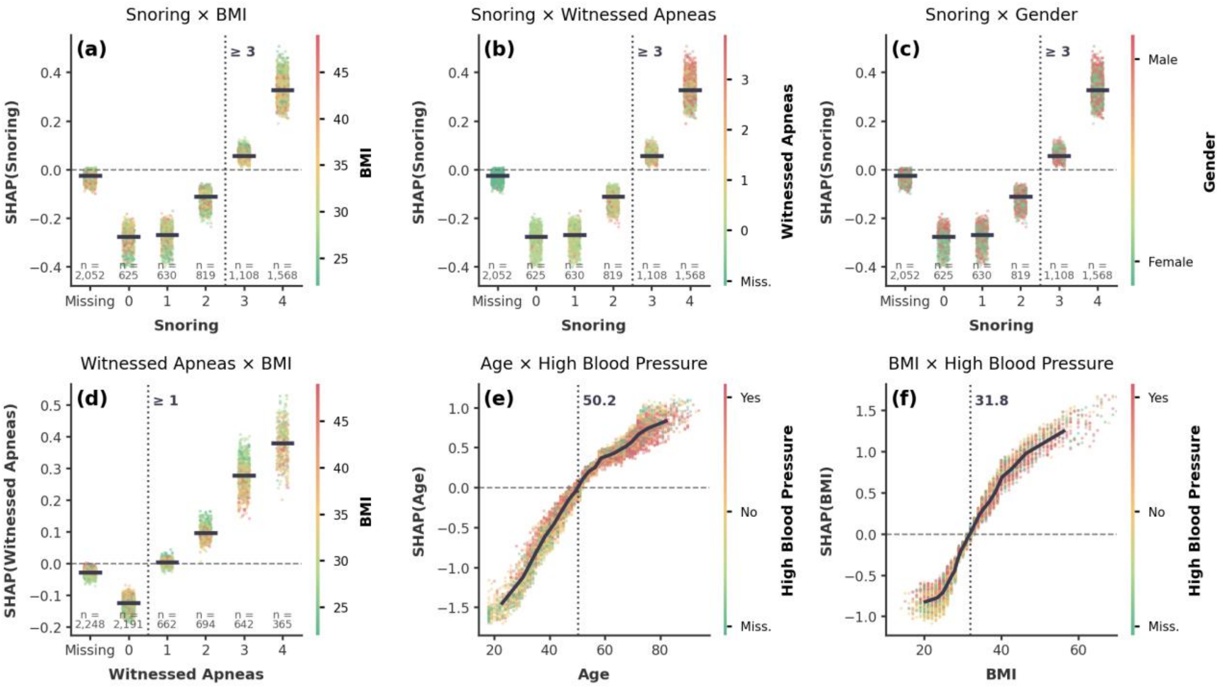


Dashed line: SHAP = 0. Solid line: mean SHAP in 20 equal-count bins. Bars: median SHAP per category, with n.
Dotted vertical line: where mean (or category median) SHAP turns positive, i.e. risk-raising.

### Supplementary Tables

#### Table S1. Cohort Data Flow

*Number of records remaining and excluded at each step between the source questionnaire database and the analytical cohort (see Figure S1). Percentages are of the 540,941 source records. AHI, apnea–hypopnea index; BMI, body mass index; PSG, polysomnography; RDI, respiratory disturbance index.*

| Step | Stage | Rule applied | Remaining, n | Excluded, n | % of source | Exclusion breakdown |
|---|---|---|---|---|---|---|
| 0 | Raw questionnaire records | All records in the source questionnaire database | 540,941 | – | 100.0 | |
| 1 | Baseline diagnostic PSG | Not a baseline diagnostic PSG (other study types) | 291,666 | 249,275 | 53.9 | • CPAP Trial: 138,495<br>• Split Study Ordered: 36,585<br>• Intervention: 31,182<br>• BiLevel Trial: 14,820<br>• other types: 28,193 |
| 2 | Plausible age and BMI | Implausible age (< 17 y) or BMI (outside 10-150 kg/m2) | 274,294 | 17,372 | 50.7 | |
| 3 | Respiratory PSG results available | Respiratory PSG results (AHI and RDI) not available: no sleep-study report | 74,296 | 199,998 | 13.7 | • Studies before 2014: 192,705 (84,071 also lack an AHI in the questionnaire)<br>• Studies from 2014 on: 7,293 |
| 4 | No BiPAP/oxygen use, AHI and RDI obtained | Respiratory-device use, or AHI and RDI not available | 72,221 | 2,075 | 13.3 | • BiPAP or oxygen use: 844<br>• CPAP use only: 6<br>• AHI and RDI not available (no respiratory-events table in the report): 1,225 |
| 5 | Analytical cohort (adults) | Age < 18 y (adults only) | 67,397 | 4,824 | 12.5 | |

#### Table S2. AUROC Uncertainty and Majority-Class Baselines

*XGBoost AUROC with 95% confidence intervals from the DeLong method. Intervals for AUROC differences are paired DeLong intervals on the same test patients for both models. The All − Demographics interval excluded zero at all six endpoints (lower bounds 0.010 to 0.028), and the All − Questionnaire interval excluded zero at all six endpoints (lower bounds 0.050 to 0.063). The retained share of above-chance AUROC (AUROC − 0.5, relative to All-Features) was 0.86–0.94 for Demographics-Only and 0.72–0.76 for Questionnaire-Only. Majority-class accuracy is the full-cohort proportion of the more frequent class; the TabPFN column gives the 5-fold cross-validated accuracy from Table S12 and its difference from that baseline. For the four-class task, the majority-class accuracy is 0.341 (AHI) and 0.339 (RDI), against an equal-class expectation of 0.25; a constant majority-class predictor would have macro-F1 of 0.127 (AHI) and 0.127 (RDI).*

| Endpoint | All-Features AUROC (95% CI) | Demographics-Only AUROC (95% CI) | Questionnaire-Only AUROC (95% CI) | All − Demographics (paired DeLong 95% CI) | All − Questionnaire (paired DeLong 95% CI) | Majority-class accuracy | TabPFN accuracy (gain over majority) |
|---|---|---|---|---|---|---|---|
| AHI ≥5 | 0.764 (0.752–0.776) | 0.749 (0.736–0.761) | 0.690 (0.677–0.704) | 0.016 (0.010–0.021) | 0.074 (0.063–0.085) | 0.662 | 0.743 (+0.081) |
| AHI ≥15 | 0.743 (0.731–0.755) | 0.718 (0.705–0.730) | 0.674 (0.661–0.687) | 0.025 (0.018–0.032) | 0.069 (0.058–0.080) | 0.679 | 0.718 (+0.039) |
| AHI ≥30 | 0.763 (0.748–0.779) | 0.726 (0.709–0.742) | 0.699 (0.682–0.717) | 0.038 (0.028–0.047) | 0.064 (0.050–0.078) | 0.858 | 0.862 (+0.004) |
| RDI ≥5 | 0.763 (0.749–0.777) | 0.746 (0.731–0.761) | 0.696 (0.680–0.711) | 0.017 (0.011–0.024) | 0.067 (0.055–0.079) | 0.797 | 0.821 (+0.024) |
| RDI ≥15 | 0.746 (0.734–0.757) | 0.723 (0.712–0.735) | 0.679 (0.666–0.691) | 0.022 (0.016–0.028) | 0.067 (0.057–0.077) | 0.542 | 0.675 (+0.133) |
| RDI ≥30 | 0.753 (0.740–0.767) | 0.717 (0.702–0.731) | 0.691 (0.676–0.707) | 0.036 (0.028–0.045) | 0.062 (0.050–0.073) | 0.794 | 0.805 (+0.011) |

#### Table S3. Exact Confidence Intervals for Sensitivity, Specificity and Precision

*Clopper–Pearson exact 95% confidence intervals for the XGBoost values in Table S7 at the fixed 0.50 threshold, using the exact numbers of cases (sensitivity), controls (specificity) and predicted positives (precision) in each endpoint's test set. Point estimates match Table S7.*

| Feature set | Endpoint | Sensitivity (95% CI) | Specificity (95% CI) | Precision (95% CI) |
|---|---|---|---|---|
| All Features | AHI ≥5 | 0.742 (0.729–0.755) | 0.646 (0.626–0.666) | 0.807 (0.794–0.819) |
| All Features | AHI ≥15 | 0.697 (0.678–0.717) | 0.658 (0.644–0.671) | 0.490 (0.473–0.508) |
| All Features | AHI ≥30 | 0.702 (0.672–0.731) | 0.687 (0.675–0.699) | 0.272 (0.255–0.290) |
| All Features | RDI ≥5 | 0.738 (0.726–0.750) | 0.646 (0.620–0.672) | 0.892 (0.883–0.901) |
| All Features | RDI ≥15 | 0.706 (0.689–0.722) | 0.637 (0.621–0.653) | 0.625 (0.609–0.641) |
| All Features | RDI ≥30 | 0.693 (0.668–0.717) | 0.676 (0.663–0.688) | 0.357 (0.339–0.375) |
| Demographics Only | AHI ≥5 | 0.733 (0.720–0.746) | 0.626 (0.605–0.646) | 0.796 (0.783–0.808) |
| Demographics Only | AHI ≥15 | 0.694 (0.674–0.714) | 0.631 (0.617–0.645) | 0.471 (0.454–0.488) |
| Demographics Only | AHI ≥30 | 0.676 (0.646–0.706) | 0.649 (0.637–0.662) | 0.243 (0.227–0.260) |
| Demographics Only | RDI ≥5 | 0.725 (0.713–0.737) | 0.633 (0.607–0.659) | 0.887 (0.877–0.896) |
| Demographics Only | RDI ≥15 | 0.703 (0.687–0.719) | 0.621 (0.605–0.637) | 0.615 (0.599–0.631) |
| Demographics Only | RDI ≥30 | 0.685 (0.660–0.710) | 0.637 (0.624–0.650) | 0.329 (0.312–0.347) |

| Feature set | Endpoint | Sensitivity (95% CI) | Specificity (95% CI) | Precision (95% CI) |
|---|---|---|---|---|
| Questionnaire Only | AHI ≥5 | 0.657 (0.643–0.670) | 0.615 (0.594–0.635) | 0.772 (0.759–0.785) |
| Questionnaire Only | AHI ≥15 | 0.616 (0.595–0.637) | 0.626 (0.612–0.640) | 0.438 (0.420–0.455) |
| Questionnaire Only | AHI ≥30 | 0.619 (0.588–0.650) | 0.665 (0.652–0.677) | 0.236 (0.219–0.253) |
| Questionnaire Only | RDI ≥5 | 0.665 (0.653–0.678) | 0.623 (0.597–0.649) | 0.875 (0.864–0.885) |
| Questionnaire Only | RDI ≥15 | 0.640 (0.623–0.657) | 0.615 (0.599–0.631) | 0.588 (0.572–0.605) |
| Questionnaire Only | RDI ≥30 | 0.615 (0.589–0.641) | 0.652 (0.639–0.665) | 0.315 (0.298–0.333) |

#### Table S4. Cohort Characteristics

*Cohort characteristics clustered by the OSA severity levels defined by the AHI values [5, 15, 30]. For continuous features, mean, standard deviation, median and interquartile range is presented, for binary count and percentages, for ordinal count and percentages.*

| Variable | Category / Statistic | Overall (N=67,397) | Normal (N=22,754) | Mild (N=22,998) | Moderate (N=12,072) | Severe (N=9,573) |
|---|---|---|---|---|---|---|
| **Age (years)** | | | | | | |
| | Mean ± SD | 54.3 ± 15.6 | 48.6 ± 16.3 | 55.9 ± 14.5 | 57.9 ± 14.1 | 59.1 ± 14.1 |
| | Median [IQR] | 55.0 [43.0–66.0] | 48.0 [36.0–61.0] | 57.0 [46.0–67.0] | 59.0 [48.0–68.0] | 61.0 [49.0–70.0] |
| **Gender** | | | | | | |
| | Female | 35,932 (53.3%) | 14,172 (62.3%) | 12,628 (54.9%) | 5,630 (46.6%) | 3,502 (36.6%) |
| | Male | 31,465 (46.7%) | 8,582 (37.7%) | 10,370 (45.1%) | 6,442 (53.4%) | 6,071 (63.4%) |
| **Race / Ethnicity** | | | | | | |
| | White (Caucasian) | 48,425 (71.9%) | 16,190 (71.2%) | 16,382 (71.2%) | 8,707 (72.1%) | 7,146 (74.6%) |
| | African American | 15,844 (23.5%) | 5,429 (23.9%) | 5,551 (24.1%) | 2,829 (23.4%) | 2,035 (21.3%) |
| | Hispanic | 2,170 (3.2%) | 769 (3.4%) | 743 (3.2%) | 379 (3.1%) | 279 (2.9%) |
| | American Indian | 372 (0.6%) | 113 (0.5%) | 147 (0.6%) | 67 (0.6%) | 45 (0.5%) |
| | Pacific Islander | 120 (0.2%) | 47 (0.2%) | 37 (0.2%) | 24 (0.2%) | 12 (0.1%) |
| | Asian | 448 (0.7%) | 200 (0.9%) | 132 (0.6%) | 63 (0.5%) | 53 (0.6%) |
| | Alaska Native | 18 (0.0%) | 6 (0.0%) | 6 (0.0%) | 3 (0.0%) | 3 (0.0%) |
| | Native Hawaiian | 0 (0.0%) | 0 (0.0%) | 0 (0.0%) | 0 (0.0%) | 0 (0.0%) |
| **BMI (kg/m²)** | | | | | | |
| | Mean ± SD | 33.2 ± 8.4 | 30.9 ± 7.8 | 33.6 ± 8.1 | 34.4 ± 8.4 | 36.6 ± 9.2 |
| | Median [IQR] | 32.0 [27.0–38.0] | 29.0 [25.0–35.0] | 32.0 [28.0–38.0] | 33.0 [29.0–39.0] | 35.0 [30.0–41.0] |
| Obese (BMI ≥ 30) | n (% non-missing) | 42,570 (63.2%) | 11,363 (49.9%) | 15,199 (66.1%) | 8,504 (70.4%) | 7,504 (78.4%) |
| Female Age > 50 | n (% non-missing) | 22,027 (32.7%) | 6,532 (28.7%) | 8,604 (37.4%) | 4,170 (34.5%) | 2,721 (28.4%) |
| **Snoring** | | | | | | |

| | | | | | | |
|---|---|---|---|---|---|---|
| | Not at all / None | 6,248 (13.2%) | 2,652 (16.9%) | 1,958 (12.0%) | 966 (11.1%) | 672 (9.9%) |
| | Slight / Few times | 6,295 (13.3%) | 2,615 (16.6%) | 2,083 (12.8%) | 934 (10.8%) | 663 (9.7%) |
| | Moderate / Sometimes | 8,185 (17.2%) | 2,921 (18.6%) | 2,892 (17.8%) | 1,371 (15.8%) | 1,001 (14.7%) |
| | Often | 11,083 (23.3%) | 3,446 (21.9%) | 3,948 (24.2%) | 2,107 (24.3%) | 1,582 (23.2%) |
| | Severe / Always | 15,681 (33.0%) | 4,085 (26.0%) | 5,410 (33.2%) | 3,299 (38.0%) | 2,887 (42.4%) |
| | Missing (n) | 19905 | 7035 | 6707 | 3395 | 2768 |
| **Witnessed Apneas — Stop Breathing** | | | | | | |
| | Not at all / None | 21,911 (48.1%) | 8,324 (55.1%) | 7,570 (48.4%) | 3,608 (43.7%) | 2,409 (37.0%) |
| | Slight / Few times | 6,615 (14.5%) | 2,209 (14.6%) | 2,335 (14.9%) | 1,169 (14.1%) | 902 (13.9%) |
| | Moderate / Sometimes | 6,942 (15.2%) | 2,126 (14.1%) | 2,410 (15.4%) | 1,351 (16.3%) | 1,055 (16.2%) |
| | Often | 6,424 (14.1%) | 1,646 (10.9%) | 2,190 (14.0%) | 1,335 (16.2%) | 1,253 (19.3%) |
| | Severe / Always | 3,646 (8.0%) | 814 (5.4%) | 1,142 (7.3%) | 801 (9.7%) | 889 (13.7%) |
| | Missing (n) | 21859 | 7635 | 7351 | 3808 | 3065 |
| **Witnessed Apneas — Gasping** | | | | | | |
| | Not at all / None | 27,192 (56.7%) | 9,339 (58.7%) | 9,439 (57.3%) | 4,824 (55.1%) | 3,590 (52.5%) |
| | Slight / Few times | 8,504 (17.7%) | 2,874 (18.1%) | 2,918 (17.7%) | 1,582 (18.1%) | 1,130 (16.5%) |
| | Moderate / Sometimes | 6,028 (12.6%) | 1,856 (11.7%) | 2,129 (12.9%) | 1,118 (12.8%) | 925 (13.5%) |
| | Often | 4,131 (8.6%) | 1,274 (8.0%) | 1,381 (8.4%) | 773 (8.8%) | 703 (10.3%) |
| | Severe / Always | 2,128 (4.4%) | 575 (3.6%) | 618 (3.7%) | 451 (5.2%) | 484 (7.1%) |
| | Missing (n) | 19414 | 6836 | 6513 | 3324 | 2741 |
| **Sleep Restless** | | | | | | |
| | Not at all / None | 9,657 (20.0%) | 2,903 (18.1%) | 3,301 (19.9%) | 1,828 (20.8%) | 1,625 (23.5%) |
| | Slight / Few times | 11,288 (23.3%) | 3,490 (21.8%) | 3,964 (23.8%) | 2,181 (24.8%) | 1,653 (23.9%) |
| | Moderate / Sometimes | 9,984 (20.6%) | 3,271 (20.4%) | 3,460 (20.8%) | 1,883 (21.4%) | 1,370 (19.8%) |
| | Often | 10,634 (22.0%) | 3,771 (23.5%) | 3,669 (22.1%) | 1,791 (20.3%) | 1,403 (20.3%) |
| | Severe / Always | 6,814 (14.1%) | 2,609 (16.3%) | 2,230 (13.4%) | 1,123 (12.8%) | 852 (12.3%) |
| | Missing (n) | 19020 | 6710 | 6374 | 3266 | 2670 |
| **Problem Going to Sleep at Night** | | | | | | |
| | Not at all / None | 12,066 (24.6%) | 3,530 (21.8%) | 4,228 (25.1%) | 2,312 (25.9%) | 1,996 (28.6%) |

| | | | | | | |
|---|---|---|---|---|---|---|
| | Slight / Few times | 10,699 (21.8%) | 3,399 (21.0%) | 3,667 (21.8%) | 2,063 (23.1%) | 1,570 (22.5%) |
| | Moderate / Sometimes | 10,789 (22.0%) | 3,491 (21.5%) | 3,713 (22.0%) | 2,046 (22.9%) | 1,539 (22.0%) |
| | Often | 9,493 (19.4%) | 3,462 (21.4%) | 3,243 (19.2%) | 1,590 (17.8%) | 1,198 (17.1%) |
| | Severe / Always | 5,931 (12.1%) | 2,323 (14.3%) | 2,005 (11.9%) | 918 (10.3%) | 685 (9.8%) |
| | Missing (n) | 18419 | 6549 | 6142 | 3143 | 2585 |
| **Grind Teeth** | | | | | | |
| | Not at all / None | 30,624 (63.4%) | 9,394 (58.8%) | 10,616 (63.9%) | 5,850 (66.4%) | 4,764 (69.2%) |
| | Slight / Few times | 6,550 (13.6%) | 2,342 (14.6%) | 2,211 (13.3%) | 1,165 (13.2%) | 832 (12.1%) |
| | Moderate / Sometimes | 4,874 (10.1%) | 1,771 (11.1%) | 1,699 (10.2%) | 785 (8.9%) | 619 (9.0%) |
| | Often | 3,581 (7.4%) | 1,344 (8.4%) | 1,247 (7.5%) | 595 (6.8%) | 395 (5.7%) |
| | Severe / Always | 2,663 (5.5%) | 1,138 (7.1%) | 841 (5.1%) | 409 (4.6%) | 275 (4.0%) |
| | Missing (n) | 19105 | 6765 | 6384 | 3268 | 2688 |
| High Blood Pressure | n (% non-missing) | 28,563 (53.1%) | 7,169 (39.9%) | 10,249 (55.4%) | 5,974 (61.0%) | 5,171 (68.3%) |
| Before Bed Sleep Aid | n (% non-missing) | 16,518 (24.5%) | 6,227 (27.4%) | 5,704 (24.8%) | 2,710 (22.4%) | 1,877 (19.6%) |

#### Table S5. QUADAS-2 Risk of Bias Assessment

*Risk of bias assessment of studies applying ML-based questionnaires or clinical data for OSA screening, evaluated using a QUADAS-2–adapted framework. Risk of bias was assessed across four domains: patient selection, index test, reference standard, and temporal alignment. Each domain was rated as low risk, some concerns, or high risk. Overall risk is the highest domain rating. PSG, polysomnography; AASM, American Academy of Sleep Medicine; AHI, apnea–hypopnea index.*

| | | **Patient Selection** | | **Index Test** | | **Reference Standard** | | **Temporal Alignment** | | |
|---|---|---|---|---|---|---|---|---|---|---|
| **Study** | **N** | **Description** | **Risk** | **Description** | **Risk** | **Description** | **Risk** | **Description** | **Risk** | **Overall** |
| Üstün et al. (2016) | 1922 | Consecutive sleep-lab patients ≥18 with broad criteria → likely representative, minimal exclusions | Low | Lasso, Ridge, Elastic Net, SVM, SLIM. | Low | Polysomnography scored by certified technologists with AASM guidelines. | Low | Index test and PSG. performed in close temporal proximity | Low | Low |
| Wu et al. (2016) | 150 | Retrospective review of outpatients; potential selection bias from small, possibly unrepresentative sample. | High | Neural fuzzy evaluation, stepwise regression, BPNNs, SVM; minimal metrics, only one AHI range. | High | Polysomnography scored by certified technologists with AASM guidelines. | Low | Index test and PSG performed in the same clinical encounter. | Low | High |

| | | | | | | | | | | |
|---|---|---|---|---|---|---|---|---|---|---|
| Huang et al. (2020) | 6875 | Excluded non-Chinese patients; some concern about generalizability beyond that population. | High | SVM, two-stage forward stepwise selection process, cross-validation; only one AHI range. | Some concerns | 2007 AASM criteria. | Low | Index test and PSG performed in the same clinical encounter. | Low | High |
| Kim et al. (2021) | 279 | Recruited from only two centers, possibly not fully representative. Small sample size. | High | LR, SVM, RF, XGBoost; permutation feature importance; train–test split; minimal metrics, only one AHI range. | Some concerns | Polysomnography scored by certified technologists per AASM guidelines. | Low | Index test and PSG performed in the same clinical encounter. | Low | High |
| Ramesh et al. (2021) | 940 patients (1,479 records) | Wisconsin Sleep Cohort with minimal exclusions → likely representative. | Low | Ensemble and traditional methods; hyperparameter tuning; minimal metrics, only one AHI range. | Some concerns | Conventional criteria (AHI-based). | Low | EHR and PSG data were linked at the patient level. | Low | Some concerns |
| Tsai et al. (2022) | 10391 | Adults 18–90 undergoing PSG for OSA → broad inclusion, minimal exclusions. | Low | LR, kNN, NB, RF, SVM; grid search; good metrics, only one AHI range; anthropometric features. | High | Overnight PSG scored per AASM 2017. | Low | Index test and PSG performed in the same clinical encounter. | Low | High |
| Holfinger et al. (2022) | 17448 | Multi-center SAGIC data → large sample, well-defined selection. | Low | ANN, RF, kernel SVM; moderate and severe AHI ranges; good metrics reported. | Low | AASM criteria. | Low | Retrospective data appropriately split; prospective cohorts independently tested. | Low | Low |
| Han & Oh (2023) | 4014 | Samsung Medical Center with minimal exclusions → representative sample. | Low | XGBoost, LightGBM, CatBoost, Random Forest, unsupervised clustering; minimal metrics, all AHI ranges. | High | AASM criteria. | Low | Index test and PSG performed in the same clinical encounter. | Low | High |

| | | | | | | | | | | |
|---|---|---|---|---|---|---|---|---|---|---|
| Huo et al. (2023) | 2886 | Sleep Heart Health Study + validated external Wisconsin cohort → robust, large population. | Low | BASH-GN machine learning–derived questionnaire. | Some concerns | AASM criteria. | Low | Index test and PSG performed in the same clinical encounter. | Low | Some concerns |
| Maniaci et al. (2023) | 498 | Inclusion criteria clearly defined, but small single-center sample → potential selection bias. | High | SVM and logistic regression; mild and severe AHI ranges. | High | AASM criteria. | Low | Index test and PSG were performed in the same clinical encounter. | Low | High |
| Liu et al. (2024) | 439 (3,390 after SMOTE oversampling) | Single center with broad inclusion → likely. representative | Low | LightGBM, RF, SVM; good metrics; moderate and severe AHI ranges. | Low | AASM criteria. | Low | Index test and PSG performed in the same clinical encounter. | Low | Low |
| Russo et al. (2025) | 420 | Excluded but single-center approach. | Some concerns | LR, classification tree, RF, XGBoost, SVM. | Low | AASM criteria. | Low | Index test and PSG performed in the same clinical encounter. | Low | Some concerns |

#### Table S6. Moses–Littenberg sROC Model Parameters

*Moses–Littenberg model parameter estimates for sROC analyses at AHI thresholds ≥5, ≥15, and ≥30 events/h, estimated by unweighted least squares from the study cohorts in Figure S6 (values from manuscript Table 2; k, number of cohorts). The intercept (α) and slope (β) describe the relationship between diagnostic log odds ratio and the accuracy–threshold parameter (S). Diagnostic odds ratios (DOR) are reported at the mean value of S and at the Q-point (S = 0).*

| Threshold | α (Intercept) | β (Slope) | DOR at Mean S | Q-point DOR (S = 0) |
|---|---|---|---|---|
| AHI ≥5 (k = 4) | 2.484 | −0.351 | 13.571 | 11.988 |
| AHI ≥15 (k = 6) | 1.766 | −0.287 | 5.450 | 5.846 |
| AHI ≥30 (k = 3) | 1.674 | −0.133 | 4.836 | 5.331 |

#### Table S7. XGBoost Binary Classification Performance — All Feature Sets

*Predictive performance of XGBoost models for binary classification across feature sets and diagnostic thresholds in the held-out test set (fixed decision threshold 0.50). Within each endpoint the three feature sets were evaluated on the same test patients (6,763–6,802 per endpoint). F1 is computed for the positive class and equals the harmonic mean of precision and sensitivity. AUROC, area under the receiver operating characteristic curve; AHI, apnea–hypopnea index; RDI, respiratory disturbance index.*

| Feature Set | Metric | AHI ≥5 | AHI ≥15 | AHI ≥30 | RDI ≥5 | RDI ≥15 | RDI ≥30 |
|---|---|---|---|---|---|---|---|
| **All Features (n=169)** | AUROC | 0.764 | 0.743 | 0.763 | 0.763 | 0.746 | 0.753 |
| | Sensitivity | 0.742 | 0.697 | 0.702 | 0.738 | 0.706 | 0.693 |
| | Specificity | 0.646 | 0.658 | 0.687 | 0.646 | 0.637 | 0.676 |
| | Precision | 0.807 | 0.490 | 0.272 | 0.892 | 0.625 | 0.357 |
| | F1 Score | 0.773 | 0.576 | 0.392 | 0.808 | 0.663 | 0.471 |
| **Demographics Only (n=4)** | AUROC | 0.749 | 0.718 | 0.726 | 0.746 | 0.723 | 0.717 |
| | Sensitivity | 0.733 | 0.694 | 0.676 | 0.725 | 0.703 | 0.685 |
| | Specificity | 0.626 | 0.631 | 0.649 | 0.633 | 0.621 | 0.637 |
| | Precision | 0.796 | 0.471 | 0.243 | 0.887 | 0.615 | 0.329 |
| | F1 Score | 0.763 | 0.561 | 0.358 | 0.798 | 0.656 | 0.445 |
| **Questionnaire Only (n=161)** | AUROC | 0.690 | 0.674 | 0.699 | 0.696 | 0.679 | 0.692 |
| | Sensitivity | 0.657 | 0.616 | 0.619 | 0.665 | 0.640 | 0.615 |
| | Specificity | 0.615 | 0.626 | 0.665 | 0.623 | 0.615 | 0.652 |
| | Precision | 0.772 | 0.438 | 0.236 | 0.875 | 0.588 | 0.315 |
| | F1 Score | 0.710 | 0.512 | 0.341 | 0.756 | 0.613 | 0.417 |

#### Table S8. Paired net-benefit differences at anchor thresholds

Difference in net benefit between feature sets on the held-out test set, with patient-level stratified bootstrap 95% percentile intervals (1000 resamples). Positive values favour the first-named feature set. Provisional, see Supplementary Methods.

| Endpoint | Contrast | Threshold | Δ net benefit | 95% CI lo | 95% CI hi |
|---|---|---|---|---|---|
| AHI ≥5 | All-Features minus Demographics-Only | 0.10 | 0.0012 | 0.0002 | 0.0023 |
| AHI ≥5 | All-Features minus Demographics-Only | 0.20 | 0.0024 | -0.0003 | 0.0053 |
| AHI ≥5 | All-Features minus Demographics-Only | 0.30 | 0.0017 | -0.0025 | 0.0057 |
| AHI ≥5 | All-Features minus Questionnaire-Only | 0.10 | 0.0008 | -0.0006 | 0.0020 |
| AHI ≥5 | All-Features minus Questionnaire-Only | 0.20 | 0.0025 | -0.0013 | 0.0061 |
| AHI ≥5 | All-Features minus Questionnaire-Only | 0.30 | 0.0045 | -0.0019 | 0.0101 |
| AHI ≥15 | All-Features minus Demographics-Only | 0.10 | 0.0027 | 0.0017 | 0.0037 |
| AHI ≥15 | All-Features minus Demographics-Only | 0.20 | 0.0097 | 0.0072 | 0.0123 |
| AHI ≥15 | All-Features minus Demographics-Only | 0.30 | 0.0179 | 0.0135 | 0.0222 |

| | | | | | |
|---|---|---|---|---|---|
| AHI ≥15 | All-Features minus Questionnaire-Only | 0.10 | 0.0036 | 0.0026 | 0.0047 |
| AHI ≥15 | All-Features minus Questionnaire-Only | 0.20 | 0.0182 | 0.0151 | 0.0211 |
| AHI ≥15 | All-Features minus Questionnaire-Only | 0.30 | 0.0368 | 0.0313 | 0.0423 |
| AHI ≥30 | All-Features minus Demographics-Only | 0.10 | 0.0045 | 0.0035 | 0.0055 |
| AHI ≥30 | All-Features minus Demographics-Only | 0.20 | 0.0167 | 0.0144 | 0.0191 |
| AHI ≥30 | All-Features minus Demographics-Only | 0.30 | 0.0318 | 0.0275 | 0.0359 |
| AHI ≥30 | All-Features minus Questionnaire-Only | 0.10 | 0.0071 | 0.0060 | 0.0081 |
| AHI ≥30 | All-Features minus Questionnaire-Only | 0.20 | 0.0298 | 0.0271 | 0.0325 |
| AHI ≥30 | All-Features minus Questionnaire-Only | 0.30 | 0.0591 | 0.0542 | 0.0642 |
| RDI ≥5 | All-Features minus Demographics-Only | 0.10 | -0.0006 | -0.0018 | 0.0007 |
| RDI ≥5 | All-Features minus Demographics-Only | 0.20 | -0.0005 | -0.0034 | 0.0025 |
| RDI ≥5 | All-Features minus Demographics-Only | 0.30 | -0.0011 | -0.0058 | 0.0034 |
| RDI ≥5 | All-Features minus Questionnaire-Only | 0.10 | -0.0016 | -0.0031 | -0.0002 |
| RDI ≥5 | All-Features minus Questionnaire-Only | 0.20 | -0.0063 | -0.0096 | -0.0028 |
| RDI ≥5 | All-Features minus Questionnaire-Only | 0.30 | -0.0129 | -0.0195 | -0.0061 |
| RDI ≥15 | All-Features minus Demographics-Only | 0.10 | 0.0013 | 0.0004 | 0.0023 |
| RDI ≥15 | All-Features minus Demographics-Only | 0.20 | 0.0050 | 0.0024 | 0.0076 |
| RDI ≥15 | All-Features minus Demographics-Only | 0.30 | 0.0067 | 0.0025 | 0.0111 |
| RDI ≥15 | All-Features minus Questionnaire-Only | 0.10 | 0.0025 | 0.0014 | 0.0034 |
| RDI ≥15 | All-Features minus Questionnaire-Only | 0.20 | 0.0099 | 0.0071 | 0.0129 |

| | | | | | |
|---|---|---|---|---|---|
| RDI ≥15 | All-Features minus Questionnaire-Only | 0.30 | 0.0223 | 0.0168 | 0.0278 |
| RDI ≥30 | All-Features minus Demographics-Only | 0.10 | 0.0040 | 0.0031 | 0.0049 |
| RDI ≥30 | All-Features minus Demographics-Only | 0.20 | 0.0122 | 0.0095 | 0.0151 |
| RDI ≥30 | All-Features minus Demographics-Only | 0.30 | 0.0258 | 0.0215 | 0.0301 |
| RDI ≥30 | All-Features minus Questionnaire-Only | 0.10 | 0.0052 | 0.0042 | 0.0061 |
| RDI ≥30 | All-Features minus Questionnaire-Only | 0.20 | 0.0206 | 0.0179 | 0.0235 |
| RDI ≥30 | All-Features minus Questionnaire-Only | 0.30 | 0.0412 | 0.0360 | 0.0467 |

#### Table S9. Calibration of the XGBoost test-set probabilities

Prevalence and mean predicted probability on the held-out test set. Calibration slope and intercept come from a logistic recalibration of the outcome on logit(probability); slope 1 and intercept 0 indicate calibration. Brier score is the mean squared error of the probabilities. Provisional, see Supplementary Methods.

| Endpoint | Feature set | Prevalence | Mean predicted | Slope | Intercept | Brier |
|---|---|---|---|---|---|---|
| AHI ≥5 | All-Features | 0.665 | 0.537 | 1.030 | 0.673 | 0.194 |
| AHI ≥5 | Demographics-Only | 0.665 | 0.529 | 1.030 | 0.692 | 0.201 |
| AHI ≥5 | Questionnaire-Only | 0.665 | 0.522 | 1.053 | 0.669 | 0.220 |
| AHI ≥15 | All-Features | 0.321 | 0.456 | 0.981 | -0.692 | 0.204 |
| AHI ≥15 | Demographics-Only | 0.321 | 0.473 | 1.014 | -0.742 | 0.215 |
| AHI ≥15 | Questionnaire-Only | 0.321 | 0.474 | 0.994 | -0.711 | 0.224 |
| AHI ≥30 | All-Features | 0.143 | 0.409 | 0.966 | -1.718 | 0.192 |
| AHI ≥30 | Demographics-Only | 0.143 | 0.435 | 0.930 | -1.761 | 0.210 |
| AHI ≥30 | Questionnaire-Only | 0.143 | 0.441 | 1.028 | -1.716 | 0.211 |
| RDI ≥5 | All-Features | 0.798 | 0.568 | 1.040 | 1.344 | 0.191 |
| RDI ≥5 | Demographics-Only | 0.798 | 0.554 | 1.040 | 1.373 | 0.200 |
| RDI ≥5 | Questionnaire-Only | 0.798 | 0.543 | 1.054 | 1.334 | 0.215 |
| RDI ≥15 | All-Features | 0.462 | 0.491 | 1.040 | -0.146 | 0.204 |
| RDI ≥15 | Demographics-Only | 0.462 | 0.493 | 1.023 | -0.147 | 0.212 |
| RDI ≥15 | Questionnaire-Only | 0.462 | 0.493 | 1.022 | -0.139 | 0.225 |
| RDI ≥30 | All-Features | 0.206 | 0.432 | 0.959 | -1.300 | 0.199 |
| RDI ≥30 | Demographics-Only | 0.206 | 0.452 | 0.938 | -1.329 | 0.214 |
| RDI ≥30 | Questionnaire-Only | 0.206 | 0.452 | 0.944 | -1.288 | 0.216 |

#### Table S10. XGBoost Four-Class Classification Performance

*Macro-averaged F1 scores and accuracy for XGBoost four-class severity classification (None, Mild, Moderate, Severe) across feature sets and indices, computed from the held-out test-set predictions; predictions were assigned to the class with the highest predicted probability. The All-Features values correspond to the confusion matrices in Figure S9. Accuracy does not exceed 0.45 in any evaluated configuration.*

| Feature Set | AHI Macro-F1 | AHI Accuracy | RDI Macro-F1 | RDI Accuracy |
|---|---|---|---|---|
| All Features (n=169) | 0.378 | 0.446 | 0.399 | 0.417 |
| Demographics Only (n=4) | 0.350 | 0.439 | 0.375 | 0.400 |
| Questionnaire Only (n=161) | 0.323 | 0.395 | 0.341 | 0.370 |

#### Table S11. SHAP Values — All-Features XGBoost Model (Four-Class Classification)

*SHAP feature importance for XGBoost four-class severity classification using all feature sets, for AHI and RDI indices. Class aggregation is the mean of the per-class mean |SHAP| values across the four classes. Rows are sorted by the AHI column; this order is not the RDI ranking. SHAP, SHapley Additive exPlanations; AHI, apnea–hypopnea index; RDI, respiratory disturbance index.*

| Feature | AHI 4-Class | RDI 4-Class |
|---|---|---|
| BMI | 0.2710 | 0.2390 |
| Age | 0.2660 | 0.2650 |
| Gender | 0.2110 | 0.2030 |
| Snoring | 0.1060 | 0.0910 |
| Witnessed Apneas | 0.0810 | 0.0650 |
| High Blood Pressure | 0.0590 | 0.0530 |
| Have Difficulty Falling Asleep | 0.0540 | 0.0510 |
| Race | 0.0350 | 0.0290 |
| Feeling Refreshing | 0.0340 | 0.0240 |
| Time in Bed (Weekday) | 0.0340 | 0.0210 |
| Sleepiness Affect Performance | 0.0330 | 0.0220 |
| Ep. Score | 0.0330 | 0.0140 |
| Sleep Get Going | 0.0330 | 0.0190 |
| Time in Bed (Weekend) | 0.0330 | 0.0190 |
| Sleep Throat Burn | 0.0320 | 0.0250 |
| Problem with Tiredness | 0.0290 | 0.0120 |
| Falling Asleep Thought | 0.0280 | 0.0170 |
| Sleep Alert | 0.0280 | 0.0080 |
| Feel Sleepy Now | 0.0280 | 0.0150 |
| Sleep Thirst | 0.0270 | 0.0180 |
| Female Age >50 | 0.0270 | 0.0220 |
| Falling Asleep Discomfort | 0.0260 | 0.0110 |
| Sleep Nasal Congestion | 0.0260 | 0.0150 |
| Sleep Restless | 0.0260 | 0.0160 |
| Problem Going to Sleep at Night | 0.0260 | 0.0100 |
| Feel Tired Now | 0.0260 | 0.0240 |
| Sleep Quality Last Night | 0.0250 | 0.0280 |
| Shift 1st | 0.0250 | 0.0210 |
| Sleep Asthma | 0.0250 | 0.0190 |
| Before Bed Sleep Aid | 0.0250 | 0.0240 |
| Someone in Room | 0.0250 | 0.0140 |
| Gasping Episodes | 0.0250 | 0.0110 |
| Feeling Tired | 0.0240 | 0.0120 |
| Grind Teeth | 0.0240 | 0.0110 |
| Weight Gain | 0.0240 | 0.0080 |
| Leg Pain While Falling Asleep | 0.0240 | 0.0130 |

| | | |
|---|---|---|
| Sleep Leg Cramps | 0.0230 | 0.0120 |
| Problem Waking Up During Night | 0.0230 | 0.0120 |
| Falling Asleep Worry | 0.0230 | 0.0140 |
| Problem with Sleepiness | 0.0220 | 0.0120 |
| Diabetes | 0.0220 | 0.0200 |
| Nasal Medication | 0.0220 | 0.0080 |
| Falling Asleep Depressed | 0.0220 | 0.0110 |
| Sleep Parasthesia | 0.0220 | 0.0130 |
| Leg Jerk | 0.0210 | 0.0140 |
| Someone in Bed | 0.0210 | 0.0150 |
| Sleep Sweat | 0.0210 | 0.0080 |
| Nap | 0.0200 | 0.0130 |
| Sleep Other Movements | 0.0200 | 0.0110 |
| Sleep Disturb Partner | 0.0200 | 0.0120 |
| Problem Not Feeling Rested | 0.0200 | 0.0100 |
| Falling Asleep Muscular Tension | 0.0200 | 0.0090 |
| Feeling Sleepy | 0.0200 | 0.0110 |
| Falling Asleep Jerk | 0.0190 | 0.0080 |
| Sleep Urinate | 0.0190 | 0.0090 |
| Sleep Talk | 0.0190 | 0.0100 |
| Sleep Special Surface | 0.0190 | 0.0090 |
| Ep. Talking | 0.0180 | 0.0120 |
| More Than One Pillow | 0.0180 | 0.0130 |
| Sleep Short of Breath | 0.0180 | 0.0090 |
| Alcohol | 0.0180 | 0.0150 |
| Feeling Alert | 0.0180 | 0.0090 |
| Falling Asleep Dream | 0.0170 | 0.0090 |
| Sleep Provide Assistance | 0.0170 | 0.0070 |
| Ep. Sitting Quietly After Lunch | 0.0170 | 0.0050 |
| Sleep Eat | 0.0170 | 0.0060 |
| Sleep Breath Problem | 0.0170 | 0.0060 |
| Ep. Sitting Quietly | 0.0170 | 0.0060 |
| Sleep Choke | 0.0170 | 0.0110 |
| Ep. In Car | 0.0170 | 0.0100 |
| Caffeinated Beverage Today | 0.0170 | 0.0050 |
| Ep. Reading | 0.0170 | 0.0080 |
| Falling Asleep Afraid | 0.0160 | 0.0090 |
| Sleep Indigestion | 0.0160 | 0.0150 |
| Sleep Hunger | 0.0160 | 0.0060 |
| Shortness of Breath | 0.0160 | 0.0060 |
| Feel Alert Now | 0.0160 | 0.0120 |
| Medications Today | 0.0160 | 0.0170 |
| Dentures | 0.0160 | 0.0120 |
| Watch TV Before Bed | 0.0150 | 0.0040 |
| Ep. Lying Down | 0.0150 | 0.0060 |
| Ep. Car Passenger | 0.0150 | 0.0060 |
| Memory Loss | 0.0150 | 0.0090 |
| Sleep Itch | 0.0150 | 0.0080 |
| Hernia | 0.0150 | 0.0060 |
| Obese | 0.0150 | 0.0150 |
| Sleep Stomach Pain | 0.0140 | 0.0080 |
| Ep. Watching TV | 0.0140 | 0.0100 |
| Wake During Night | 0.0140 | 0.0150 |
| Naps Today | 0.0140 | 0.0090 |
| Physical Complaints Today | 0.0140 | 0.0090 |
| Headache | 0.0140 | 0.0160 |
| Difficulty Going Back to Sleep | 0.0140 | 0.0110 |
| Sleep Chest Pain | 0.0130 | 0.0050 |

| | | |
|---|---|---|
| Frequent Cough | 0.0130 | 0.0050 |
| Depression | 0.0130 | 0.0110 |
| Sleep Cough | 0.0130 | 0.0070 |
| Second Awaken Part | 0.0130 | 0.0050 |
| Breathing Difficulty | 0.0130 | 0.0080 |
| Swollen Legs | 0.0130 | 0.0070 |
| Sleep Heart Pain | 0.0130 | 0.0040 |
| First Awaken Part | 0.0130 | 0.0060 |
| Awake Feeling | 0.0130 | 0.0070 |
| Joint Pain | 0.0120 | 0.0050 |
| Nasal Discharge | 0.0120 | 0.0080 |
| Tobacco Habit | 0.0120 | 0.0040 |
| Irregular Heart Beat | 0.0120 | 0.0040 |
| Other Heart Disease | 0.0110 | 0.0050 |
| Slept Longer | 0.0110 | 0.0100 |
| Falling Asleep Paralyzed | 0.0110 | 0.0040 |
| Cancer | 0.0100 | 0.0050 |
| Work Sleep Fatigue | 0.0100 | 0.0050 |
| Weight Loss | 0.0100 | 0.0050 |
| Too Little Sleep | 0.0100 | 0.0040 |
| Heartburn | 0.0100 | 0.0050 |
| Feeling Sick | 0.0100 | 0.0050 |
| Allergy | 0.0100 | 0.0030 |
| Feel Sleepy Today | 0.0100 | 0.0030 |
| Voice Changes | 0.0100 | 0.0050 |
| History Tobacco Use | 0.0100 | 0.0030 |
| Dreaming Last Night | 0.0090 | 0.0070 |
| Arthritis | 0.0090 | 0.0070 |
| Body Shaking | 0.0090 | 0.0040 |
| Sleep Walk | 0.0090 | 0.0050 |
| Muscular Pain | 0.0090 | 0.0030 |
| Swallowing Difficulty | 0.0080 | 0.0060 |
| Caffeinated Beverage | 0.0080 | 0.0040 |
| Alcohol Quit | 0.0080 | 0.0040 |
| Kidney Disease | 0.0080 | 0.0040 |
| Third Awaken Part | 0.0070 | 0.0050 |
| Chest Pain | 0.0070 | 0.0040 |
| Thyroid Disease | 0.0070 | 0.0030 |
| Sleep Difficulty | 0.0070 | 0.0040 |
| Stroke History | 0.0060 | 0.0030 |
| Enlarged Heart | 0.0060 | 0.0030 |
| Muscle Weakness | 0.0060 | 0.0040 |
| Painful Sinus | 0.0060 | 0.0040 |
| Stomach Disease | 0.0060 | 0.0040 |
| Heart Attack | 0.0060 | 0.0030 |
| Medication Past Month | 0.0050 | 0.0050 |
| Peptic Ulcer | 0.0050 | 0.0020 |
| Smothering Spells at Night | 0.0050 | 0.0020 |
| Personality Changes | 0.0050 | 0.0040 |
| Anemia | 0.0050 | 0.0020 |
| Physical Strenuous Day | 0.0050 | 0.0020 |
| Out of Ordinary Happen Today | 0.0040 | 0.0030 |
| Body Control Difficulty | 0.0040 | 0.0010 |
| Low Blood Pressure | 0.0040 | 0.0020 |
| Enlarged Thyroid | 0.0040 | 0.0020 |
| Seizures | 0.0040 | 0.0030 |
| Liver Disease | 0.0040 | 0.0030 |
| Too Much Sleep | 0.0040 | 0.0020 |

| | | |
|---|---|---|
| Sleep Related Misses | 0.0040 | 0.0020 |
| Paralysis Attacks | 0.0040 | 0.0020 |
| Speaking Difficulty | 0.0040 | 0.0030 |
| Alcohol Beverage Today | 0.0040 | 0.0040 |
| Tuberculosis | 0.0030 | 0.0020 |
| Abnormal Glucose | 0.0030 | 0.0020 |
| Low Blood Sugar | 0.0030 | 0.0020 |
| Normal Weight | 0.0030 | 0.0150 |
| Shift 3rd | 0.0030 | 0.0010 |
| Shift 2nd | 0.0030 | 0.0010 |
| Intestinal Disease | 0.0030 | 0.0030 |
| Sleep Related Accidents | 0.0030 | 0.0020 |
| Sleep Accident Fatigue | 0.0030 | 0.0010 |
| HIV | 0.0030 | 0.0010 |
| Sleep Accident Mistakes | 0.0020 | 0.0010 |
| Shift Swing | 0.0010 | 0.0010 |
| Underweight | 0.0000 | 0.0000 |

#### Table S12. TabPFN Binary Classification Performance

*TabPFN binary classification performance for AHI and RDI severity thresholds based on 5-fold cross-validation, with a fixed decision threshold of 0.50. Values are mean ± standard deviation across folds. Lower thresholds (≥5) prioritise sensitivity; higher thresholds (≥30) demonstrate high specificity at the expense of recall. AHI, apnea–hypopnea index; RDI, respiratory disturbance index.*

| Index | Threshold | AUROC | Accuracy | Sensitivity | Specificity | Precision | F1 Score |
|---|---|---|---|---|---|---|---|
| AHI | ≥5 | 0.762 ± 0.003 | 0.743 ± 0.003 | 0.901 ± 0.004 | 0.433 ± 0.005 | 0.757 ± 0.005 | 0.823 ± 0.003 |
| AHI | ≥15 | 0.743 ± 0.006 | 0.718 ± 0.004 | 0.341 ± 0.006 | 0.896 ± 0.006 | 0.608 ± 0.013 | 0.437 ± 0.005 |
| AHI | ≥30 | 0.773 ± 0.005 | 0.862 ± 0.002 | 0.096 ± 0.010 | 0.989 ± 0.001 | 0.590 ± 0.049 | 0.166 ± 0.016 |
| RDI | ≥5 | 0.763 ± 0.004 | 0.821 ± 0.005 | 0.963 ± 0.002 | 0.264 ± 0.006 | 0.837 ± 0.006 | 0.895 ± 0.003 |
| RDI | ≥15 | 0.742 ± 0.004 | 0.675 ± 0.005 | 0.637 ± 0.007 | 0.708 ± 0.004 | 0.648 ± 0.005 | 0.643 ± 0.006 |
| RDI | ≥30 | 0.759 ± 0.006 | 0.805 ± 0.003 | 0.166 ± 0.005 | 0.970 ± 0.003 | 0.592 ± 0.027 | 0.259 ± 0.008 |

#### Table S13. Concentration of Mean |SHAP| in the Top-Ranked Features

*Share of the summed mean |SHAP| over all features that is carried by the three highest-ranked features (and by the highest-ranked feature) in the XGBoost binary models, computed from the mean |SHAP| values underlying Table S14 (rounded to three decimals, so shares are approximate). Age, BMI and gender are the top three features in the All-Features models. Spearman rank correlation of the 169 mean |SHAP| values between endpoints was 0.61–0.86 (All-Features; 15 endpoint pairs) and 0.64–0.91 (Questionnaire-Only; 161 features); the AHI-threshold pairs gave 0.63, 0.73 and 0.81 for All-Features (≥5 vs ≥30, ≥5 vs ≥15, ≥15 vs ≥30).*

| Feature set | Endpoint | Three highest-ranked features | Top-3 share of summed mean \|SHAP\| (%) | Top-1 share (%) |
|---|---|---|---|---|
| All Features | AHI ≥5 | Age, BMI, Gender | 51.1 | 19.6 |
| All Features | AHI ≥15 | Age, BMI, Gender | 40.5 | 14.4 |
| All Features | AHI ≥30 | BMI, Age, Gender | 39.1 | 14.1 |
| All Features | RDI ≥5 | Age, BMI, Gender | 45.0 | 17.0 |
| All Features | RDI ≥15 | Age, BMI, Gender | 42.3 | 15.7 |

| Feature set | Endpoint | Three highest-ranked features | Top-3 share of summed mean \|SHAP\| (%) | Top-1 share (%) |
|---|---|---|---|---|
| All Features | RDI ≥30 | BMI, Age, Gender | 34.0 | 12.1 |
| Questionnaire Only | AHI ≥5 | High Blood Pressure, Snoring, Witnessed Apneas | 20.0 | 8.7 |
| Questionnaire Only | AHI ≥15 | High Blood Pressure, Snoring, Witnessed Apneas | 17.4 | 6.6 |
| Questionnaire Only | AHI ≥30 | High Blood Pressure, Witnessed Apneas, Snoring | 17.4 | 7.3 |
| Questionnaire Only | RDI ≥5 | High Blood Pressure, Snoring, Witnessed Apneas | 19.4 | 8.5 |
| Questionnaire Only | RDI ≥15 | High Blood Pressure, Snoring, Witnessed Apneas | 15.9 | 6.1 |
| Questionnaire Only | RDI ≥30 | High Blood Pressure, Have Difficulty Falling Asleep, Snoring | 14.9 | 6.1 |

#### Table S14. SHAP Values — All-Features XGBoost Model (138 of 169 Features)

*Mean |SHAP| for the All-Features XGBoost binary model at AHI thresholds ≥5, ≥15 and ≥30 events/h, ranked by mean |SHAP| at AHI ≥5. The 138 features with the highest AHI ≥5 values are shown; the remaining 31 features (ranks 139–169) have mean |SHAP| ≤ 0.001 at AHI ≥5 and are omitted. Demographic variables (age, BMI, gender) dominate across all thresholds. SHAP, SHapley Additive exPlanations; AHI, apnea–hypopnea index.*

| Rank | Feature | AHI ≥ 5 | AHI ≥ 15 | AHI ≥ 30 |
|---|---|---|---|---|
| 1 | Age | 0.5270 | 0.4550 | 0.4810 |
| 2 | BMI | 0.4970 | 0.4480 | 0.5430 |
| 3 | Gender | 0.3510 | 0.3780 | 0.4760 |
| 4 | Snoring | 0.1610 | 0.1680 | 0.1670 |
| 5 | Witnessed Apneas | 0.1070 | 0.1230 | 0.1490 |
| 6 | High Blood Pressure | 0.0920 | 0.0980 | 0.1290 |
| 7 | Before Bed Sleep Aid | 0.0290 | 0.0350 | 0.0320 |
| 8 | Obese | 0.0230 | 0.0080 | 0.0020 |
| 9 | Feel Sleepy Now | 0.0230 | 0.0170 | 0.0140 |
| 10 | Diabetes | 0.0220 | 0.0320 | 0.0350 |
| 11 | Sleep Asthma | 0.0200 | 0.0270 | 0.0400 |
| 12 | Someone in Room | 0.0200 | 0.0230 | 0.0310 |
| 13 | Depression | 0.0200 | 0.0070 | 0.0030 |
| 14 | Sleep Get Going | 0.0180 | 0.0120 | 0.0140 |
| 15 | Sleepiness Affect Performance | 0.0170 | 0.0300 | 0.0480 |
| 16 | Sleep Throat Burn | 0.0170 | 0.0390 | 0.0440 |
| 17 | Sleep Other Movements | 0.0160 | 0.0160 | 0.0080 |
| 18 | Ep. Score | 0.0160 | 0.0230 | 0.0290 |
| 19 | Dentures | 0.0160 | 0.0150 | 0.0170 |
| 20 | Sleep Restless | 0.0160 | 0.0210 | 0.0240 |
| 21 | Sleep Sweat | 0.0160 | 0.0080 | 0.0100 |
| 22 | Problem Not Feeling Rested | 0.0150 | 0.0200 | 0.0140 |
| 23 | Feeling Sleepy | 0.0150 | 0.0050 | 0.0050 |
| 24 | Sleep Talk | 0.0140 | 0.0080 | 0.0010 |
| 25 | Problem with Sleepiness | 0.0140 | 0.0090 | 0.0020 |
| 26 | Physical Complaints Today | 0.0130 | 0.0130 | 0.0110 |
| 27 | Leg Jerk | 0.0130 | 0.0130 | 0.0090 |
| 28 | Normal Weight | 0.0130 | 0.0100 | 0.0060 |

| | | | | |
|---|---|---|---|---|
| 29 | Alcohol | 0.0130 | 0.0150 | 0.0010 |
| 30 | Female Age >50 | 0.0120 | 0.0090 | 0.0070 |
| 31 | Grind Teeth | 0.0120 | 0.0210 | 0.0230 |
| 32 | Wake During Night | 0.0120 | 0.0130 | 0.0190 |
| 33 | Sleep Thirst | 0.0110 | 0.0230 | 0.0190 |
| 34 | Falling Asleep Worry | 0.0110 | 0.0090 | 0.0120 |
| 35 | Problem with Tiredness | 0.0110 | 0.0110 | 0.0260 |
| 36 | Falling Asleep Thought | 0.0110 | 0.0290 | 0.0380 |
| 37 | Falling Asleep Muscular Tension | 0.0110 | 0.0080 | 0.0150 |
| 38 | Race | 0.0100 | 0.0360 | 0.0720 |
| 39 | More Than One Pillow | 0.0100 | 0.0080 | 0.0040 |
| 40 | Swollen Legs | 0.0100 | 0.0080 | 0.0190 |
| 41 | Problem Going to Sleep at Night | 0.0100 | 0.0130 | 0.0070 |
| 42 | Falling Asleep Afraid | 0.0100 | 0.0090 | 0.0070 |
| 43 | Ep. Car Passenger | 0.0100 | 0.0040 | 0.0060 |
| 44 | Caffeinated Beverage Today | 0.0090 | 0.0030 | 0.0090 |
| 45 | Joint Pain | 0.0090 | 0.0020 | 0.0050 |
| 46 | Have Difficulty Falling Asleep | 0.0090 | 0.0920 | 0.1440 |
| 47 | Ep. Talking | 0.0090 | 0.0170 | 0.0200 |
| 48 | Weight Gain | 0.0080 | 0.0210 | 0.0260 |
| 49 | Headache | 0.0080 | 0.0230 | 0.0320 |
| 50 | Feel Tired Now | 0.0080 | 0.0230 | 0.0490 |
| 51 | Falling Asleep Discomfort | 0.0080 | 0.0060 | 0.0310 |
| 52 | Sleep Nasal Congestion | 0.0080 | 0.0210 | 0.0120 |
| 53 | Sleep Short of Breath | 0.0080 | 0.0040 | 0.0030 |
| 54 | Frequent Cough | 0.0080 | 0.0070 | 0.0020 |
| 55 | Other Heart Disease | 0.0080 | 0.0030 | 0.0070 |
| 56 | Breathing Difficulty | 0.0070 | 0.0140 | 0.0290 |
| 57 | Ep. Sitting Quietly | 0.0070 | 0.0100 | 0.0120 |
| 58 | Time in Bed (Weekend) | 0.0070 | 0.0190 | 0.0360 |
| 59 | Falling Asleep Jerk | 0.0070 | 0.0070 | 0.0060 |
| 60 | Sleep Disturb Partner | 0.0070 | 0.0060 | 0.0020 |
| 61 | Feeling Tired | 0.0070 | 0.0120 | 0.0200 |
| 62 | Tobacco Habit | 0.0070 | 0.0030 | 0.0010 |
| 63 | Someone in Bed | 0.0060 | 0.0130 | 0.0310 |
| 64 | Sleep Chest Pain | 0.0060 | 0.0060 | 0.0070 |
| 65 | Leg Pain While Falling Asleep | 0.0060 | 0.0080 | 0.0050 |
| 66 | Falling Asleep Dream | 0.0060 | 0.0050 | 0.0020 |
| 67 | Ep. Reading | 0.0050 | 0.0060 | 0.0090 |
| 68 | Problem Waking Up During Night | 0.0050 | 0.0190 | 0.0060 |
| 69 | Time in Bed (Weekday) | 0.0050 | 0.0170 | 0.0370 |
| 70 | Sleep Cough | 0.0050 | 0.0050 | 0.0040 |
| 71 | Feeling Alert | 0.0050 | 0.0060 | 0.0070 |
| 72 | Memory Loss | 0.0050 | 0.0110 | 0.0140 |
| 73 | Sleep Choke | 0.0050 | 0.0130 | 0.0040 |
| 74 | Sleep Leg Cramps | 0.0050 | 0.0100 | 0.0120 |
| 75 | Sleep Indigestion | 0.0050 | 0.0070 | 0.0020 |
| 76 | Falling Asleep Depressed | 0.0050 | 0.0040 | 0.0080 |
| 77 | Feeling Refreshing | 0.0050 | 0.0130 | 0.0440 |
| 78 | Hernia | 0.0050 | 0.0140 | 0.0160 |
| 79 | Cancer | 0.0050 | 0.0020 | 0.0050 |
| 80 | Gasping Episodes | 0.0040 | 0.0160 | 0.0100 |
| 81 | Sleep Urinate | 0.0040 | 0.0110 | 0.0150 |
| 82 | Ep. Sitting Quietly After Lunch | 0.0040 | 0.0060 | 0.0110 |
| 83 | Sleep Alert | 0.0040 | 0.0150 | 0.0100 |
| 84 | Low Blood Pressure | 0.0040 | 0.0020 | 0.0020 |

| | | | | |
|---|---|---|---|---|
| 85 | Caffeinated Beverage | 0.0040 | 0.0030 | 0.0020 |
| 86 | Ep. Lying Down | 0.0040 | 0.0050 | 0.0020 |
| 87 | Feel Alert Now | 0.0040 | 0.0060 | 0.0050 |
| 88 | Nasal Medication | 0.0040 | 0.0060 | 0.0040 |
| 89 | Liver Disease | 0.0040 | 0.0010 | 0.0030 |
| 90 | Sleep Provide Assistance | 0.0040 | 0.0060 | 0.0030 |
| 91 | Slept Longer | 0.0040 | 0.0030 | 0.0040 |
| 92 | Awake Feeling | 0.0040 | 0.0030 | 0.0150 |
| 93 | Sleep Special Surface | 0.0030 | 0.0060 | 0.0040 |
| 94 | Ep. Watching TV | 0.0030 | 0.0030 | 0.0230 |
| 95 | Dreaming Last Night | 0.0030 | 0.0020 | 0.0020 |
| 96 | Body Shaking | 0.0030 | 0.0040 | 0.0000 |
| 97 | Weight Loss | 0.0030 | 0.0100 | 0.0190 |
| 98 | Enlarged Heart | 0.0030 | 0.0030 | 0.0020 |
| 99 | Sleep Hunger | 0.0030 | 0.0030 | 0.0020 |
| 100 | Naps Today | 0.0030 | 0.0150 | 0.0330 |
| 101 | Feeling Sick | 0.0030 | 0.0010 | 0.0050 |
| 102 | Sleep Parasthesia | 0.0030 | 0.0160 | 0.0380 |
| 103 | Heartburn | 0.0030 | 0.0050 | 0.0020 |
| 104 | Shift 1st | 0.0030 | 0.0210 | 0.0550 |
| 105 | Sleep Itch | 0.0030 | 0.0060 | 0.0070 |
| 106 | mh_tobacco_history | 0.0030 | 0.0030 | 0.0010 |
| 107 | Feel Sleepy Today | 0.0020 | 0.0040 | 0.0000 |
| 108 | Sleep Heart Pain | 0.0020 | 0.0030 | 0.0020 |
| 109 | Sleep Walk | 0.0020 | 0.0020 | 0.0010 |
| 110 | Sleep Quality Last Night | 0.0020 | 0.0350 | 0.0770 |
| 111 | Too Little Sleep | 0.0020 | 0.0100 | 0.0010 |
| 112 | Nap | 0.0020 | 0.0250 | 0.0290 |
| 113 | Chest Pain | 0.0020 | 0.0030 | 0.0000 |
| 114 | Sleep Breath Problem | 0.0020 | 0.0040 | 0.0010 |
| 115 | Heart Attack | 0.0020 | 0.0030 | 0.0010 |
| 116 | Peptic Ulcer | 0.0020 | 0.0010 | 0.0010 |
| 117 | Medication Past Month | 0.0020 | 0.0040 | 0.0050 |
| 118 | Kidney Disease | 0.0020 | 0.0040 | 0.0100 |
| 119 | Sleep Eat | 0.0020 | 0.0040 | 0.0050 |
| 120 | Ep. In Car | 0.0020 | 0.0170 | 0.0180 |
| 121 | Smothering Spells at Night | 0.0020 | 0.0020 | 0.0010 |
| 122 | Sleep Stomach Pain | 0.0020 | 0.0040 | 0.0060 |
| 123 | Second Awaken Part | 0.0020 | 0.0030 | 0.0020 |
| 124 | Shortness of Breath | 0.0010 | 0.0070 | 0.0070 |
| 125 | Allergy | 0.0010 | 0.0070 | 0.0010 |
| 126 | Speaking Difficulty | 0.0010 | 0.0010 | 0.0010 |
| 127 | Sleep Difficulty | 0.0010 | 0.0030 | 0.0000 |
| 128 | Abnormal Glucose | 0.0010 | 0.0000 | 0.0000 |
| 129 | First Awaken Part | 0.0010 | 0.0150 | 0.0080 |
| 130 | Alcohol Beverage Today | 0.0010 | 0.0030 | 0.0000 |
| 131 | Difficulty Going Back to Sleep | 0.0010 | 0.0040 | 0.0150 |
| 132 | Irregular Heart Beat | 0.0010 | 0.0080 | 0.0080 |
| 133 | Stroke History | 0.0010 | 0.0030 | 0.0060 |
| 134 | Third Awaken Part | 0.0010 | 0.0030 | 0.0040 |
| 135 | Thyroid Disease | 0.0010 | 0.0020 | 0.0010 |
| 136 | Shift 2nd | 0.0010 | 0.0010 | 0.0010 |
| 137 | Too Much Sleep | 0.0010 | 0.0010 | 0.0010 |
| 138 | Out of Ordinary Happen Today | 0.0010 | 0.0010 | 0.0060 |

#### Supplementary Methods: decision curve analysis

Net benefit for referring patients with predicted probability at or above a threshold pt was NB(pt) = TP/n − (FP/n) × pt/(1 − pt), with treat-all and treat-none as reference strategies (Vickers and Elkin, 2006). Curves were computed on the held-out test set of each endpoint over thresholds 0.01 to 0.80 (step 0.01). Paired differences between feature sets were estimated at thresholds 0.10, 0.20 and 0.30 by a patient-level bootstrap with 1000 resamples and seed 42. Resampling was stratified by outcome, so the case count was fixed in every resample, and the same patients were used for both models in each resample. Percentile 95% intervals are reported. The three anchor thresholds were chosen for description; no clinical threshold range has been confirmed.

Calibration. The binary XGBoost probabilities were not recalibrated. Calibration slopes lay between 0.93 and 1.05, but intercepts were offset: the mean predicted probability differed from prevalence (for example 0.54 against 0.67 at AHI ≥5, and 0.41 against 0.14 at AHI ≥30; Supplementary Table S9). This pattern is consistent with the class weighting used in training. Net benefit computed on these probabilities is therefore provisional, and recalibration on out-of-fold training predictions is required before clinical interpretation. No cost data were available, so no cost-effectiveness analysis is reported.
